# Rare and *de novo* variants in 827 congenital diaphragmatic hernia probands implicate *LONP1* and *ALYREF* as new candidate risk genes

**DOI:** 10.1101/2021.06.01.21257928

**Authors:** Lu Qiao, Le Xu, Lan Yu, Julia Wynn, Rebecca Hernan, Xueya Zhou, Christiana Farkouh-Karoleski, Usha S. Krishnan, Julie Khlevner, Aliva De, Annette Zygmunt, Timothy Crombleholme, Foong-Yen Lim, Howard Needelman, Robert A. Cusick, George B. Mychaliska, Brad W. Warner, Amy J. Wagner, Melissa E. Danko, Dai Chung, Douglas Potoka, Przemyslaw Kosiński, David J. McCulley, Mahmoud Elfiky, Kenneth Azarow, Elizabeth Fialkowski, David Schindel, Samuel Z. Soffer, Jane B. Lyon, Jill M. Zalieckas, Badri N. Vardarajan, Gudrun Aspelund, Vincent P. Duron, Frances A. High, Xin Sun, Patricia K. Donahoe, Yufeng Shen, Wendy K. Chung

## Abstract

Congenital diaphragmatic hernia (CDH) is a severe congenital anomaly that is often accompanied by other anomalies. Although the role of genetics in the pathogenesis of CDH has been established, only a small number of disease genes have been identified. To further investigate the genetics of CDH, we analyzed *de novo* coding variants in 827 proband-parent trios and confirmed an overall significant enrichment of damaging *de novo* variants, especially in constrained genes. We identified *LONP1* (Lon Peptidase 1, Mitochondrial) and *ALYREF* (Aly/REF Export Factor) as novel candidate CDH genes based on *de novo* variants at a false discovery rate below 0.05. We also performed ultra-rare variant association analyses in 748 cases and 11,220 ancestry-matched population controls and identified *LONP1* as a risk gene contributing to CDH through both *de novo* and ultra-rare inherited largely heterozygous variants clustered in the core of the domains and segregating with CDH in familial cases. Approximately 3% of our CDH cohort was heterozygous with ultra-rare predicted damaging variants in *LONP1* who have a range of clinical phenotypes including other anomalies in some individuals and higher mortality and requirement for extracorporeal membrane oxygenation. Mice with lung epithelium specific deletion of *Lonp1* die immediately after birth and have reduced lung growth and branching that may at least partially explain the high mortality in humans. Our findings of both *de novo* and inherited rare variants in the same gene may have implications in the design and analysis for other genetic studies of congenital anomalies.

## Introduction

Congenital diaphragmatic hernia (CDH) affects approximately 3 per 10,000 neonates^1,2^. Approximately 40% of CDH cases occur with additional congenital anomalies besides common secondary anomalies (dextrocardia and lung hypoplasia)^3^. The most common additional anomalies^4,5^ are structural heart defects (11-15%), musculoskeletal malformations (15-20%) including limb deficiency, club foot, and omphalocele^6^. However, anomalies of almost every organ have been described in association with CDH. Despite advances in care including improved prenatal diagnosis, fetal interventions, extracorporeal membrane oxygenation (ECMO) and gentle ventilation, CDH continues to be associated with at least 20% mortality and significant long-term morbidity including feeding difficulties, pulmonary hypertension and other respiratory complications, and neurocognitive deficits^3,7,8^.

The complexity of the phenotypes associated with CDH is mirrored by the complexity of the genetics, which are heterogeneous with approximately 30% of CDH cases having an identifiable major genetic contributor. Typically, each gene or copy number variant (CNV) associated with CDH accounts for at most 1-2% of cases^9^. The full spectrum of genomic variants has been associated with CDH, including chromosome aneuploidies (10%), copy number variants (CNVs) (3-10%), monogenic conditions (10-22%), and emerging evidence for oligogenic causes^1^ (CNVs and individual genes^10^).

While familial cases have been described, CDH most commonly occurs in individuals without a family history of CDH, and sibling recurrence risk in isolated cases is less than 1%^11^. Likely due to the historically high mortality and low reproductive fitness, CDH is often due to *de novo* CNVs and single gene variants. However, dominant inheritance has been described with transmission of an incompletely penetrant variant from an unaffected parent or parent with a subclinical diaphragm defect^12^. CDH has also been described in individuals with biallelic variants such as Donnai-Barrow syndrome^13^. The occurrence of discordant monogenic twins suggests a role for stochastic events after fertilization^11^.

A genetic diagnosis for probands with CDH can inform prognosis and guide medical management. Some genetic conditions associated with CDH are associated with an increased risk for additional anomalies, increased mortality, and increased morbidity including neurocognitive disabilities that may benefit from early intervention^3^. Over the past decade, advances in genomic sequencing technology have helped to define the genes associated with CDH. We and others have shown that *de novo* variants with large effect size contribute to 10-22% of CDH cases with enrichment of *de novo* likely damaging variants in CDH cases with an additional anomaly (complex CDH) ^9,14,15^. We also demonstrated a higher burden of *de novo* likely damaging (LD) variants in females compared to males supporting a “female protective model”^9^. Most recently, in a cohort with long term developmental outcome data^3^, we demonstrated that *de novo* likely damaging (LD) variants are associated with poorer neurodevelopmental outcomes as well as a higher prevalence of pulmonary hypertension (PH).

To expand upon our knowledge of the diverse genetic etiologies of CDH, we performed whole genome (WGS) or exome sequencing of 827 CDH proband-parent trios. We confirmed an overall enrichment of damaging *de novo* variants in constrained genes, and identified *LONP1* (Lon Peptidase 1, Mitochondrial [MIM: 605490]) and *ALYREF* (Aly/REF Export Factor [MIM: 604171]) as new candidate CDH genes with recurrent ultra-rare and *de novo* variants.

## Materials and methods

### Participant recruitment and control datasets

Study participants were enrolled as fetuses, neonates, children and adults with a radiologically confirmed diaphragm defect by the DHREAMS study^16^ (Diaphragmatic Hernia Research & Exploration; Advancing Molecular Science) or Boston Children’s Hospital/Massachusetts General Hospital (BCH/MGH) as described previously^14^. Clinical data were prospectively collected from medical records and entered into a central Research Electronic Data Capture (REDCap) database^17^. Probands and both parents provided a blood, skin biopsy, or saliva specimen for trio genetic analysis. All studies were approved by the Columbia University institutional review board (IRB), serving as the central site. Each participating site also procured approval from their local IRB and signed informed consent was obtained. Ethical approval was obtained from the following participating institutions: Boston Children’s Hospital/Massachusetts General Hospital, Washington University, Cincinnati Children’s Hospital Medical Center, Children’s Hospital & Medical Center of Omaha, University of Michigan, Monroe Carell Jr. Children’s Hospital, Northwell Health, Oregon Health & Science University, Legacy Research Institute, University of Texas Southwestern, Children’s Hospital of Wisconsin, and Children’s Hospital of Pittsburgh.

A total of 827 cases and their parents had whole genome (WGS) or exome sequencing in the current study. A subset of trios (n=574) has been described in our previous study^3,9^.

Participants with only a diaphragm defect were classified as isolated CDH while participants with at least one additional major congenital anomaly (*e*.*g*. congenital heart defect, central nervous system anomaly, gastrointestinal anomaly, skeletal anomaly, genitourinary anomaly, cleft lip/palate), moderate to severe developmental delay, or other neuropsychiatric phenotypes at last contact were classified as complex CDH. Pulmonary hypoplasia, cardiac displacement and intestinal herniation were considered to be part of the diaphragm defect sequence and were not considered independent malformations. Data on the child’s current and past health including family history of congenital anomalies, postoperative pulmonary hypertension, mortality or survival status prior to initial discharge, extracorporeal membrane oxygenation (ECMO) intake were gathered as described previously^3^.

The control group consisted of unaffected parents from the Simons Powering Autism Research for Knowledge (SPARK) study^18^ (exomes) and Latinx samples from Washington Heights-Hamilton Heights-Inwood Community Aging Project (WHICAP) study^19^ (exomes).

### WGS and exome data analysis

There are 233 CDH trios processed using whole genome sequencing (WGS) that were not included in previous studies^3,9^ (Table S1). Of these 233 previously unpublished trios, 1 trio was processed at Baylor College of Medicine Human Genome Sequencing Center and 232 trios at Broad Institute Genomic Services. The genomic libraries of 219 cases were prepared by TruSeq DNA PCR-Free Library Prep Kit (Illumina), while 14 were TruSeq DNA PCR-Plus Library Prep Kit (Illumina), with average fragment length about 350 bp, and sequenced as paired-end of 150-bp on Illumina HiSeq X platform. Exome sequencing was performed in 20 CDH trios that were not previously published^3,9^. Among these, the coding exons of 9 trios were captured using Agilent Sure Select Human All Exon Kit v2 (Agilent Technologies), 10 trios using NimbleGen SeqCap EZ Human Exome V3 kit (Regeneron NimbleGen), 1 trio using NimbleGen SeqCap EZ Human Exome V2 kit (Roche NimbleGen). Exomes of SPARK cohort were captured using a slightly modified version of the IDT xGen Exome Research Panel v.1.0 identical to the previous study^20^. Whole‐exome sequencing of the WHICAP cohort was performed at Columbia University using the Roche SeqCap EZ Exome Probes v3.0 Target Enrichment Probes^21^.

Exome and WGS data of cases and controls were processed using a pipeline implementing GATK Best Practice v4.0 as previously described^9,22^. Specifically, reads of exome cases were mapped to human genome GRCh37 reference using BWA-MEM^23^, while reads of WGS cases, SPARK and WHICAP controls were mapped to GRCh38; duplicated reads were marked using Picard^24^; variants were called using GATK^25^ (v4.0) HaplotypeCaller to generate gVCF files for joint genotyping. All samples within the same batch (Table S1) were jointly genotyped and variant quality score recalibration (VQSR) was performed using GATK. To combine all cases for further analysis, we lifted over the GRCh37 variants to GRCh38 using CrossMap^26^ (v0.3.0). Common SNP genotypes within exome regions were used to validate familial relationships using KING^27^ and ancestries using peddy^28^ (v0.4.3) in cases, SPARK controls and WHICAP controls.

*De novo* variants were defined as a variant present in the offspring with homozygous reference genotypes in both parents. Here, we limited WGS to coding regions based on coding sequences and canonical splice sites of all GENCODE v27 coding genes. We took a series of stringent filters to identify *de novo* variants as described previously^9^: VQSR tranche ≤99.8 for SNVs and ≤99.0 for indels; GATK’s Fisher Strand ≤25, quality by depth ≥2. We required the candidate *de novo* variants in probands to have ≥5 reads supporting the alternative allele, ≥20% alternative allele fraction, Phred-scaled genotype likelihood ≥60 (GQ), and population allele frequency ≤0.01% in gnomAD v2.1.1; both parents to have ≥10 reference reads, <5% alternative allele fraction, and GQ ≥30. We applied DeepVariant^29^ to all candidate *de novo* variants for in silico confirmation and only included the ones with PASS from DeepVariant for downstream analysis.

To reduce batch effects in combined datasets from different sources^30^ in analysis of rare variants, for non-Latinx population we targeted ultra-rare variants located in xGen-captured protein coding regions and for Latinx population in regions targeted by xGen and SeqCap EZ v3.0. We used the following criteria to minimize technical artifacts and select ultra-rare variants^22^: cohort AF <0.5% and population cohort <1×10^−5^ across all genomes in gnomAD v3.0; mappability=1; >90% target region with depth ≥10; overlapped with segmental duplication regions <95%; genotype quality >30, allele balance >20% and depth >10 in cases.

We used Ensembl Variant Effect Predictor^31^ (VEP, Ensemble 102) and ANNOVAR^32^ to annotate variant function, variant population frequencies and *in silico* predictions of deleteriousness. All coding SNVs and indels were classified as synonymous, missense, inframe, or likely-gene-disrupting (LGD, which includes frameshift indels, canonical splice site, or nonsense variants). We defined predicted damaging missense (D-mis) based on CADD^33^ score v1.3. All *de novo* variants and inherited variants in candidate risk genes were manually inspected in the Integrative Genome Viewer (IGV). A total of 179 variants were selected for validation using Sanger sequencing; all of them were confirmed (Table S2). To compare the clinical outcomes between cases with deleterious variants in candidate genes and with likely damaging (LD) variants, we defined likely damaging variants as in our previous study^3^: (a) *de novo* LGD or deleterious missense variants in genes that are constrained (ExAC pLI 0.9) and highly expressed in developing diaphragm^34^, or (b) *de novo* LGD or deleterious missense variants in known risk genes for CDH or commonly comorbid disorders (congenital heart disease [CHD] and neurodevelopmental delay [NDD]), or (c) plausible deleterious missense variants in known risk genes for CDH or commonly comorbid disorders (CHD and NDD), or (d) deletions in constrained (ExAC pLI≥0.9) or haploinsufficient genes from ClinGen genome dosage map^35^, or (e) CNVs implicated in known syndromes. We classified CDH cases into two genetic groups: (1) LD, if the case carried at least one *de novo* LD variant; (2) non-LD, if the case carried no such variants.

*De novo* copy number variants (CNVs) were identified using an inhouse pipeline of read depth-based algorithm based on CNVnator^36^ v0.3.3 in WGS trios as described in our previous study^3^. The *de novo* CNV segments were validated by the additional pair-end/split-read (PE/SR) evidence using Lumpy^37^ v0.2.13 and SVtyper^38^ v0.1.4. Only the CNVs supported by both read depth (RD) and PE/SR were included in downstream analysis. We mapped *de novo* CNVs on GENCODE v29 protein coding genes with at least 1bp in the shared interval. The GENCODE genes were annotated with variant intolerance metric by ExAC pLI^39^, haploinsufficiency metric by Episcore^40^, haploinsufficiency and triplosensitivity of genes from ClinGen genome dosage map^35^, and CNV syndromes from DECIPHER^41^ v11.1.

### Quantitative PCR

We performed experimental validation of putative *de novo* genic CNVs using quantitative PCR (qPCR). All PCR primers were designed for the selected genes located within the *de novo* CNVs and synthesized by IdtDNA. All qPCR reactions were performed in a total of 10 μl volume, comprising 5 μl 2x SYBR Green I Master Mix (Promega), 1 μl 10nM of each primer and 2 μl of 1:20 diluted cDNA in 96-well plates using CFX Connect Real-Time PCR Detection System (Bio-Rad). All reactions were performed in triplicate, and the conditions were 5 minutes at 95°C, then 40 cycles of 95°C at 15 seconds and 60°C at 30 seconds. The relative copy numbers were calculated using the standard curve method relative to the β-actin housekeeping gene. Five-serial 4-fold dilutions of DNA samples were used to construct the standard curves for each primer.

### Statistical analysis

#### Burden of de novo variants

The baseline mutation rates for different classes of *de novo* variants were calculated in each GENCODE coding gene using the published trinucleotide sequence context^42^, and we calculated the rate in protein-coding regions that are uniquely mappable as previously described mutation model^9,18^. The observed number of variants of various types (*e*.*g*. synonymous, missense, LGD) in each gene set and case group was compared with the baseline expectation using Poisson test. In all analyses, constrained genes were defined by ExAC pLI^39^ score of >0.5, and all remaining genes were treated as other genes. We used a less stringent pLI threshold than previously suggested^39^ for defining constrained genes, because it captures more known haploinsufficient genes important for heart and diaphragm development. We compared the observed number of variants in female versus male cases and complex versus isolated cases using the binormal test.

#### extTADA analysis

To identify risk genes based on *de novo* variants, we used an empirical Bayesian method, extTADA^43^ (Extended Transmission and *de novo* Association). The extTADA model was developed based on a previous integrated empirical Bayesian model TADA^44^ and estimates mean effect sizes and risk-gene proportions from the genetic data using MCMC (Markov Chain Monte Carlo) process (details see supplemental note). To inform the parameter estimation with prior knowledge of developmental disorders, we stratify the genes into constrained genes (ExAC pLI score >0.5) and non-constrained genes (other genes), followed by estimating the parameters using the extTADA model to each group of genes. After estimating posterior probability of association (PPA) of individual genes in each group, we combined both groups to calculate a final false discovery rate (FDR) for each gene using extTADA’s procedure.

#### Gene-based case-control association analysis of ultra-rare variants

To identify novel risk genes based both on *de novo* and rare inherited variants, we performed a gene-based association test comparing the frequency of ultra-rare deleterious variants in CDH cases with controls, without considering *de novo* status. Samples with read depth coverage ≥10x for 80% in exome cases and 90% in genome cases of the targeted regions were included in the analysis (Figure S1). Relatedness was checked using KING^27^, and only unrelated cases were included in the association tests (Figure S2). To control for confounding from genetic ancestry, we selected ancestry-matched controls using SPARK exomes and Latinx WHICAP exomes to reach a fixed case/control ratio in each population ancestry inferred by peddy^28^ (Figure S3). Specifically, for a specific ancestry ($), consider *x*_*i*_ number of cases, *y*_*i*_ number of controls, *n*_*i*_ the fold controls to cases (*y*_*i*_/*x*_*i*_). We chose the minimized *n*_*min*_ among all ancestries. In each genetic-ancestry group controls (*y*_*i*_), we ranked the Euclidean distance between each case and controls which were calculated from top 3 PCA eigenvectors and selected *n*_*min*_*x*_*i*_ controls from *y*_*i*_ controls to ensure the same proportions in cases and controls. After filtering to reduce the impact of false positive variants, we tested for similarity of the ultra-rare synonymous variant rate among cases and controls in specific genetic-ancestry groups, assuming that ultra-rare synonymous variants are mostly neutral with respect to disease status.

To identify CDH risk genes, we tested the burden of ultra-rare deleterious variants (AF <1×10^−5^ across all gnomAD v3.0 genomes, LGD or D-Mis) in each protein-coding gene in cases compared to controls. To improve statistical power, we searched for a gene-specific CADD^33^ score threshold for defining D-Mis that maximized the burden of ultra-rare deleterious variants in cases compared to controls and used permutations to calculate statistical significance with the variable threshold test^22,45^. For the binomial tests in each permutation, we used binom.test function in R to calculate p values. We performed two association tests, one with LGD and D-Mis variants combined and the other with D-Mis variants alone, to account for different modes of action. We defined the threshold for genome-wide significance by Bonferroni correction for multiple testing (as two tests for each gene with 20,000 protein-coding genes, threshold p-value=1.25 ×10^−6^). We checked for inflation using a quantile-quantile (Q-Q) plot and calculated the genomic control factor (lambda [λ]) using QQperm in R. Lambda equal to 1 indicates no deviation from the expected distribution.

### Protein modeling

We searched the LONP1 canonical sequence (identifier: P36776-1) in UniProt and obtained the structural model of the human mitochondrial LONP1 monomer (encompassing only the residue range 413–951) using SWISS-MODEL server^46^ with SMTL ID 6u5z.1 as template. The 3D structure was visualized using PyMOL molecular viewer (The PyMOL Molecular Graphics System, Version 1.2r3pre, Schrödinger, LLC).

### Mice

All mice were housed in American Association for Accreditation of Laboratory Animal Care accredited facilities and laboratories at University of California, San Diego. All animal experiments were conducted under approved guidelines for the Care and Use of Laboratory Animals. *Lonp1*^*fl*^ and *Shh*^*cre*^ mice have all been described previously^47^ (International Mouse Strain Resource J:204812). All mice were bred on a C57BL/6J background, and littermates were used as controls to minimize potential genetic background effects.

## Results

### Cohort characteristics

Participants were recruited as part of the multi-site DHREAMS study (n=748) and from the Boston Children’s Hospital/Massachusetts General Hospital (n=79). We performed WGS on 734 proband-parent trios and exome sequencing on 93 trios. In total, we analyzed 827 trios with WGS or exome sequencing.

In the cohort, there were 486 (59%) male probands (Table 1), consistent with a higher prevalence of CDH in males^9,48,49^. The genetically determined ancestries (Figure S3A) were European (73.4%), admixed American (hereafter referred to as Latinx; 18.5%), African (3.7%), East Asian (1.8%), and South Asian (2.5%). Among the 277 (33.5%) complex cases, the most frequent additional anomalies were congenital heart disease (n=144), NDD (n=54), skeletal anomalies (n=46), genitourinary anomalies (n=46) and gastrointestinal anomalies (n=42). A total of 533 (64.4%) probands had isolated CDH without additional anomalies at the time of last follow up. The most common type of CDH was left-sided Bochdalek (Table 1).

**Table 1.**
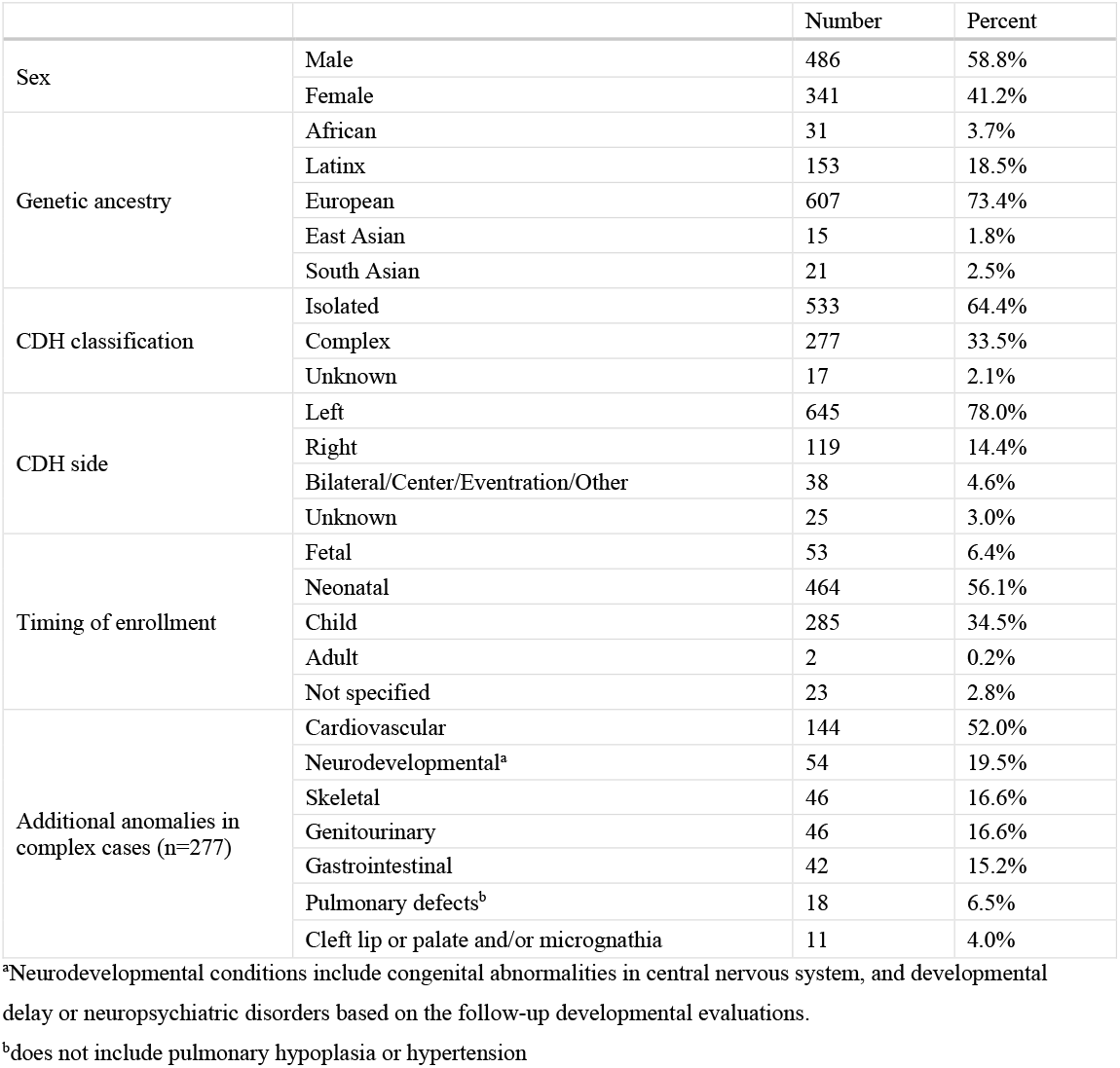
Clinical summary of 827 CDH probands.

|  |  | Number | Percent |
| --- | --- | --- | --- |
| Sex | Male | 486 | 58.8% |
|  | Female | 341 | 41.2% |
| Genetic ancestry | African | 31 | 3.7% |
|  | Latinx | 153 | 18.5% |
|  | European | 607 | 73.4% |
|  | East Asian | 15 | 1.8% |
|  | South Asian | 21 | 2.5% |
| CDH classification | Isolated | 533 | 64.4% |
|  | Complex | 277 | 33.5% |
|  | Unknown | 17 | 2.1% |
| CDH side | Left | 645 | 78.0% |
|  | Right | 119 | 14.4% |
|  | Bilateral/Center/Eventration/Other | 38 | 4.6% |
|  | Unknown | 25 | 3.0% |
| Timing of enrollment | Fetal | 53 | 6.4% |
|  | Neonatal | 464 | 56.1% |
|  | Child | 285 | 34.5% |
|  | Adult | 2 | 0.2% |
|  | Not specified | 23 | 2.8% |
| Additional anomalies in complex cases (n=277) | Cardiovascular | 144 | 52.0% |
|  | Neurodevelopmental<sup>a</sup> | 54 | 19.5% |
|  | Skeletal | 46 | 16.6% |
|  | Genitourinary | 46 | 16.6% |
|  | Gastrointestinal | 42 | 15.2% |
|  | Pulmonary defects<sup>b</sup> | 18 | 6.5% |
|  | Cleft lip or palate and/or micrognathia | 11 | 4.0% |

<sup>a</sup>Neurodevelopmental conditions include congenital abnormalities in central nervous system, and developmental delay or neuropsychiatric disorders based on the follow-up developmental evaluations.
<sup>b</sup>does not include pulmonary hypoplasia or hypertension

### Burden of *de novo* coding variants

We identified 1153 *de novo* protein-coding variants in 619 (74.8%) cases including 1058 single nucleotide variants (SNVs) and 95 indels (Table S2). The average number of *de novo* coding variants per proband is 1.39. The number of *de novo* coding variants across probands closely follows a Poisson distribution (Figure S4). Transition-to-transversion ratio of *de novo* SNVs was 2.75. We classified variants that were likely gene disruptive (LGD) or predicted damaging missense (“D-mis” with CADD score≥25) as damaging variants. A total of 418 damaging variants (126 LGD and 292 D-mis) were identified in 318 (38.4%) cases, including 83 (10%) cases harboring two or more such variants.

We analyzed the burden of *de novo* variants in CDH cases by comparing the observed number of variants to the expected number based on the background mutation rate. Consistent with previous studies on CDH^9^ and other developmental disorders^50-52^, both *de novo* LGD (0.15 per case) and D-mis variants (0.35 per case) were significantly enriched in cases (relative risk [RR]=1.5, P=3.6×10^−5^ for LGD; RR=1.3, P=3.1×10^−6^; Figures 1A and B; Table S3) while the frequency of synonymous variants (0.30 per case) closely matches the expectation (RR=0.9, P=0.12; Table S3). The burden of LGD variants is mostly located in constrained (ExAC^39^ pLI >0.5) genes (RR=2.2, P=1.8×10^−8^). It is marginally higher in female cases than male cases (RR=3.0 vs 1.36, P=0.012) and marginally higher in complex cases than isolated cases (RR=3.1 vs 1.75, P=0.024; Figure 1C; Table S3).

**Figure 1.**
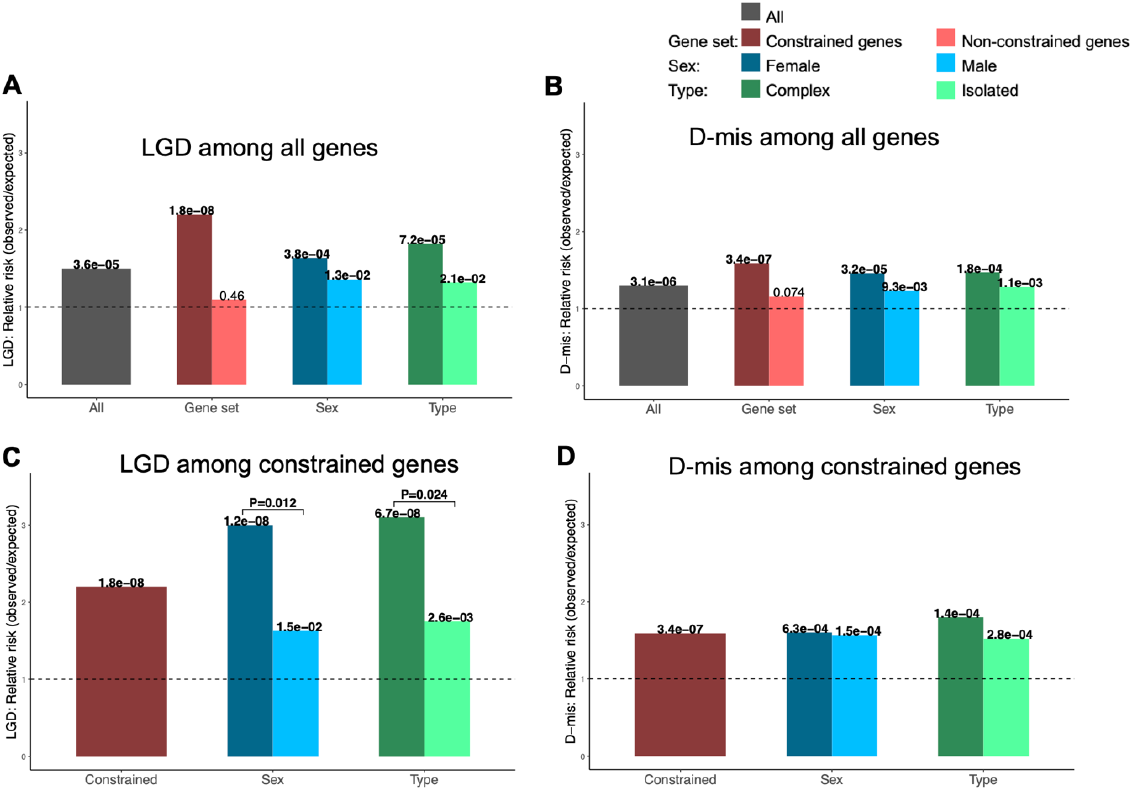
Burden of *de novo* coding variants in CDH compared to expectation. (A) LGD among all genes; (B) D-mis among all genes; (C) LGD among constrained genes; (D) D-mis among constrained genes. P values between cases and expectation by Poisson test are labeled for each bar. P values between females and males, complex and isolated cases by binormal test are labeled for each pair. Significant P values are highlighted in bold.

To identify new CDH risk genes by *de novo* variants, we applied extTADA^43^ to the data of 827 CDH trios. ExtTADA assumes a model of genetic architecture compatible with the observed burden and recurrence of *de novo* damaging variants and estimates a false discovery rate (FDR) for each gene using MCMC. From the burden analysis of *de novo* variants in CDH and previous studies^52^, we reasoned that the constrained genes (ExAC pLI >0.5) drive the higher burden of *de novo* damaging variants and are more likely to be plausible risk genes. We stratified the data into the constrained gene set and the non-constrained gene set (Table S4) and estimated extTADA priors (mean relative risk and prior probability of being a risk gene) in these two gene sets separately. Constrained genes had a higher prior of risk genes than non-constrained genes (0.037 vs 0.006). Meanwhile, both LGD and D-mis had higher relative risks in constrained genes than non-constrained gene (18.30 vs 5.24 for LGD; 10.01 vs 3.81 for D-mis). We estimated Bayes Factor of individual genes within each gene group and then combined the genes from two groups together to calculate FDR. We identified 3 genes with FDR <0.05: *MYRF* (Myelin Regulatory Factor [MIM: 608329]), *LONP*1, and *ALYREF*. Five of 6 *MYRF de novo* variants were described in our previous study^9^. We identified 3 participants harboring *de novo* D-mis variants in *LONP1* and 2 participants for *de novo* LGD variants in *ALYREF*. Of two participants with an *ALYREF* LGD variant, one had an isolated left-side CDH and the other had right-side CDH and ventricular septal defect. There were nine additional genes with ≥2 *de novo* predicted deleterious variants (*HSD17B10* [MIM: 300256], *GATA4* [MIM: 600576], *SYMPK* [MIM: 602388], *PTPN11* [MIM: 176876], *WT1* [MIM: 607102], *FAM83H* [MIM: 611927], *CACNA1H* [MIM: 607904], *SEPSECS* [MIM: 613009], and *ZFYVE26* [MIM: 612012]) (Table 2). Of these, three are known CDH genes (*MYRF, GATA4, WT1*). All *de novo* variants in these genes are heterozygous.

**Table 2.**
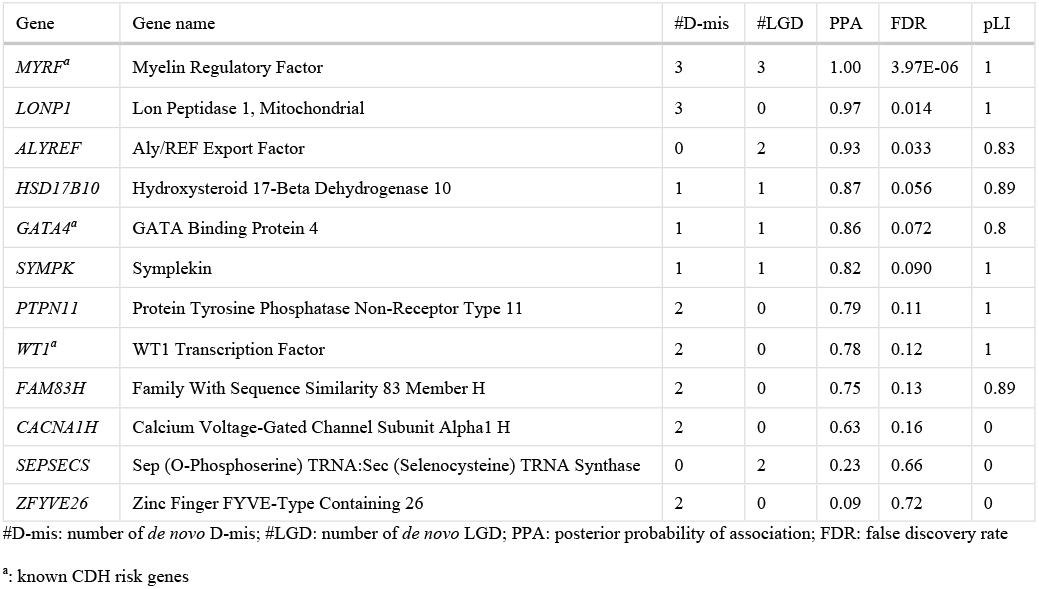
Top CDH associated genes predicted by pLI-stratified extTADA with ≥2 *de novo* predicted deleterious variant.

### Recurrent genes in *de novo* CNVs

We applied CNVnator to call CNVs from WGS data and used customized filters to identify *de novo* CNVs. We performed experimental validation of 25 putative *de novo* genic CNVs including all 9 small CNVs (<5kb) using quantitative PCR (qPCR). 22 of 25 (88%) reported *de novo* CNV in cases were confirmed by qPCR. Removing the 3 false positive CNVs, there were 87 *de novo* CNVs identified in 734 CDH cases with WGS with an average of 0.12 per case (Table S5). Among them, there were 54 (62%) deletions ranging from 2,096 bp to 33.7 Mb and 33 (38%) duplications ranging from 1,165 bp to 24.9 Mb. Seven samples carried known syndromic CNVs in DECIPHER^41^ dataset, one of which was heterozygous for a 16p13.11 microduplication, two heterozygous for a 17q12 deletion associated with renal cysts and diabetes (RCAD), three heterozygous for 21q22 duplication in the critical region for Down syndrome, and one heterozygous for 22q11 deletion associated with DiGeorge syndrome. No recurrent genes were identified between *de novo* SNVs and CNVs. Four CNVs were recurrent (Table 3), two of which encompass single genes *CSMD1* (CUB And Sushi Multiple Domains 1 [MIM: 608397]) and *GPHN* (Gephyrin [MIM: 603930]).

**Table 3.**
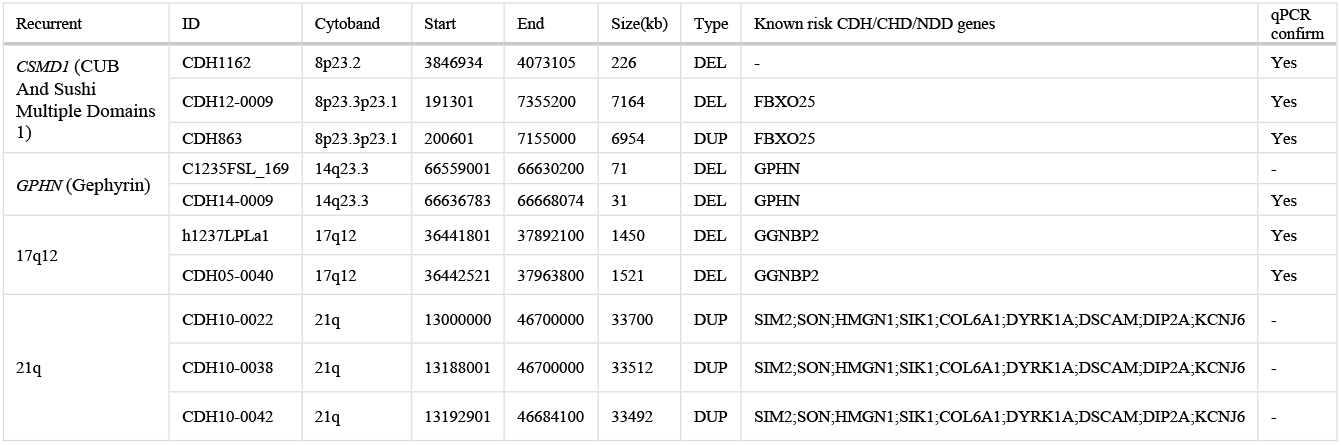
Recurrent genes or regions in *de novo* CNVs.

### Candidate gene *LONP1* contributes to CDH risk through both *de novo* and rare inherited variants

To identify additional risk genes that may contribute through rare inherited variants, we performed a gene-based, case-control association analysis of ultra-rare variants. Specifically, we used exome data from the SPARK (unaffected parents) and Latinx WHICAP samples as controls. Quality control procedures included at least 10x depth of sequence coverage across the target regions (Figure S1) and detection of cryptic relatedness amongst all CDH participants and controls (Figure S2). To prevent confounding by genetic ancestry, we performed principal component analysis (PCA) by peddy to infer genetic ancestry of all cases and controls and selected matching controls (15-fold of cases numbers in each specific genetic-ancestry group) to reach a fixed case/control ratio. With the same genetic-ancestry proportion in cases and controls (77% Europeans, 14.8% Latinx, 4.1% Africans, 2% East Asians, 2.1% South Asians; Figure S3; Table S6), we selected 748 cases and 11,220 controls for downstream analysis. We filtered the ultra-rare variant call sets of cases and controls in each genetic-ancestry group by empirical filters to reduce false positive calls and minimize technical batch effects across data sets. After filtering, the average numbers of ultra-rare (AF<1×10^−5^ across all gnomAD v3.0 genomes) synonymous variants per subject in cases and controls are nearly identical in everyone (enrichment rate=1, P=1) and specific ancestral groups (Table S7). Furthermore, a gene-level burden test confined to ultra-rare synonymous variants was consistent with a global null model in Q-Q plot (Figure S5), indicating that technical batch effects would likely have minimal impact on genetic analyses. We then performed a variable threshold association test^22,45^ to identify new risk genes based on enrichment of ultra-rare damaging variants in individual genes. For each gene, we tested enrichment of LGD and D-mis variants together or just D-mis variants, in order to account for potential different biological modes of action. In the variable threshold test, we determined a gene-specific optimal CADD score threshold to define D-mis in order to maximize the power of the association test and then estimated type I error rate by permutations. The overall result from the case-control association did not show inflation from the null model (λ=1.09; Figure 2A). The association of *LONP1* (P=1×10^−7^; Figure 2) exceeded the Bonferroni-corrected significance threshold (1.25×10^−6^, account for two tests in each gene). Three of the 24 ultra-rare deleterious variants in *LONP1* were known *de novo* variants. Two known CDH risk genes, *ZFPM2* (Zinc Finger Protein, FOG Family Member 2 [MIM: 603693]) and *MYRF*, fell just below the cutoff for genome wide significance.

**Figure 2.**
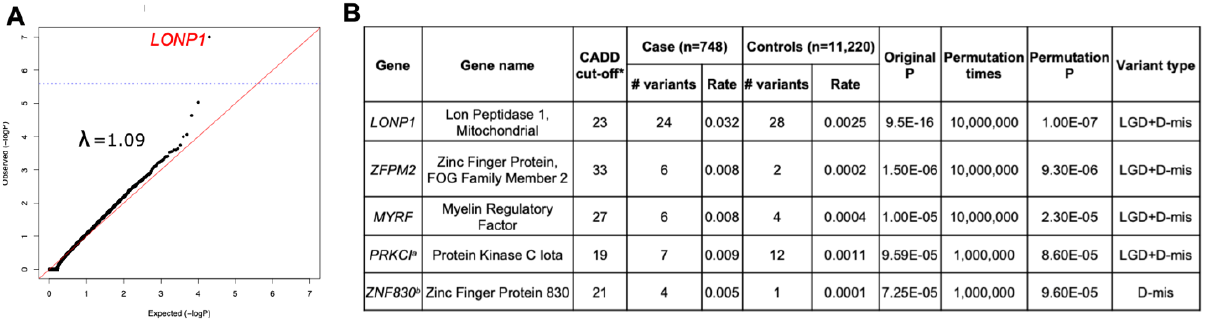
Gene-based association analysis using 748 CDH cases and 11,220 controls across all populations. (A) Results of a binomial test confined to ultra-rare LGD and D-Mis variants or D-Mis only variants in 18,939 protein-coding genes. Horizontal blue line indicates the Bonferroni-corrected threshold for significance. (B) Complete list of top association genes with permutation P values <1×10^−4^. *: a gene-specific CADD score threshold for defining D-Mis that maximized the burden of ultra-rare deleterious variants in cases compared to controls; #: numbers of deleterious variants; a: MIM 600539; b: no MIM number.

The association of *LONP1* is due to both LGD and D-mis variants. We screened the whole cohort (Figure 3 and Table 4), including CDH relatives (n=1) and exome sequencing singletons (n=2), for ultra-rare damaging missense (CADD ≥25) and LGD in *LONP1* (NM_004793.3). A total of 23 CDH cases in 829 cases (2.8%) carry 24 *LONP1* variants, including 10 LGD and 14 D-mis variants. Among 22 *LONP1* variants excluding 2 of unknown inheritance variants in singletons, there are 3 (13.6%) *de novo* variants (all D-mis) and 19 (86.4%) inherited variants, 36.8% of which are from mothers (n=7). Of 19 inherited variants, 8 parents carrying *LONP1* variants have a family history of CDH or diaphragm eventration (n=4) or other congenital anomaly (n=4; brain abnormality, cerebral palsy, cleft palate, skeletal abnormality) segregating with the *LONP1* variant. Three inherited variants (c.1913C>T [p.638M], c.2122G>A [p.G708S] and c.2263C>G [p.R755G]) are each observed twice in the cohort on different probands. Familial segregation was established in six familial CDH cases for c.398C>G (p.P133R), c.6391G>T (p.X213_splice), c.1262delG (p.F421Lfs*87), c.1574C>T (p.P525L), c.1913C>T (p.T638M) and c.2719dupG (p.V907Gfs*73). One case (01-1279) carries biallelic heterozygous variants with c.1574C>T (p.P525L) inherited from one parent and c.2263C>G (p.R755G) inherited from another parent. The participant with biallelic heterozygous variants required ECMO and died at 8-9 hours after birth with severe bilateral CDH with near complete diaphragm agenesis, bilateral lung hypoplasia, and no additional anomalies (Figure 4). All other cases are heterozygous variants.

**Figure 3.**
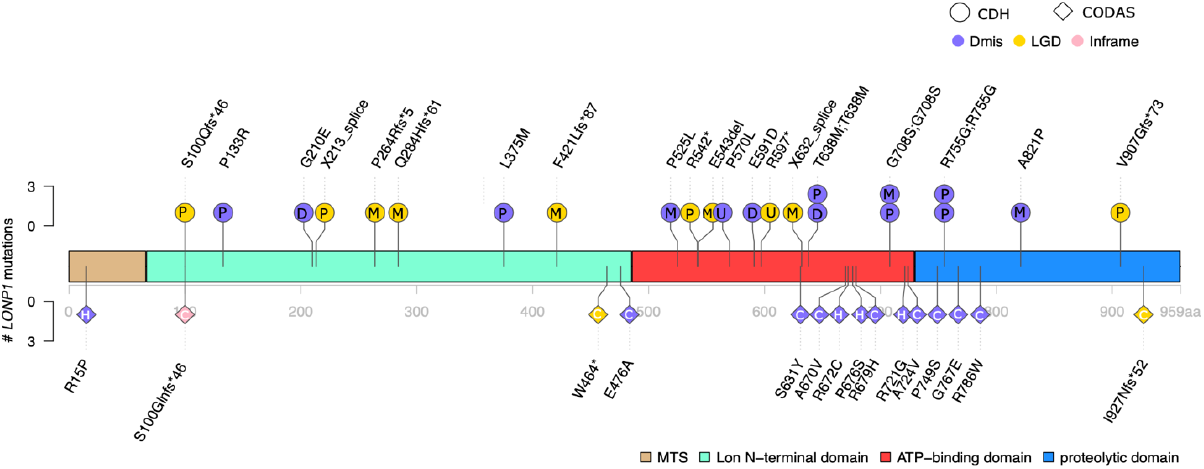
Variant locations in *LONP1* (GenBank: NM_004793.3) of CDH and CODAS syndrome. There are three main domains in LONP1, N-terminal Lon domain, ATP binding domain and proteolytic domain. Positions indicated at upper structure are variants in CDH. Deleterious heterozygous variant such as LGD and missense with CADD ≥25 and allele frequency (AF) <1e-5 across all gnomAD genomes in CDH are presented. Deleterious missense is presented in purple, LGD in yellow, inframe variant in pink. Inheritance pattern were labelled in circles of variants (P: paternal; M: maternal; D: *de novo*; U: singleton unknown). Positions at lower structure are variants in published CODAS syndrome samples. CODAS syndrome is caused by biallelic variants in *LONP1*, including homozygous (H) or compound heterozygous (C) variants in the diamonds.

**Table 4.** Phenotypes of CDH cases with ultra-rare deleterious variants in *LONP1*. Deleterious heterozygous variants include LGD and missense with CADD ≥25 with minor allele frequency (MAF) <1e-5 across all the gnomAD v3.0 genomes.

| cDNA change | Protein Change | Sample ID | Sex | Genetic ancestry | Inheritance | Family history of other birth defects | Familial CDH | 1M PH | 3M PH | Vital status | ECMO | Complex | Neuro-related | Other Congenital Anomalies/medical problems |
| --- | --- | --- | --- | --- | --- | --- | --- | --- | --- | --- | --- | --- | --- | --- |
| c.296dup | p.S100Qfs*46 | 01-0794 | Female | EUR | paternal | No | No | - | - | Deceased | No | No |  | No |
| c.398C>G | p.P133R | 01-0672 | Female | AFR | paternal | No | Affected sibling (+) | Unk | Unk | Alive | Yes | Yes | No | congenital cataracts |
| c.398C>G | p.P133R | 01-0670 | Male | AFR | paternal | No | Affected sibling (+) | - | - | Deceased | Yes | Yes |  | GI anomaly, GU anomaly |
| c.629G>A | p.G210E | 01-0070 | Male | EUR | *de novo* | No | No | - | - | Deceased | Yes | No |  | No |
| c.639-1G>T | p.X213_splice | 04-0022 | Female | EUR | paternal | Paternal half-brother with idiopathic PH (N/T) | Affected sibling (N/T), Affected paternal grandmother (+, *de novo*) | Severe | - | Deceased | Yes | No |  | No |
| c.792del | p.P264Rfs*5 | 09-0003 | Female | EUR | maternal | Maternal great uncle with suspected cerebral palsy | No | Severe | - | Deceased | Yes | No | Seizures | No |
| c.851del | p.Q284Hfs*61 | 1428 | Female | EUR | maternal | No | No | Mild | Mild | Alive | No | No | No | short stature |
| c.1123C>A | p.L375M | 04-0045 | Male | EUR | paternal | Paternal uncle: neonatal death due to brain abnormality (hydrocephalus?) | No | None | None | Alive | No | No | No | No |
| c.1262del | p.F421Lfs*87 | 1733 | Female | EUR | maternal | No | Maternal uncle with suspected CDH (N/T) | - | - | Deceased | Yes | Yes | global encephalopathy, seizures | CHD |
| c.1574C>T | p.P525L | 01-1279 | Male | EUR | maternal<sup>a</sup> | No | Affected sibling (+) | - | - | Deceased | Yes | No |  | No, bilateral CDH |
| c.1624C>T | p.R542* | 01-0113 | Male | EUR | paternal | Mother with Klippel Feil syndrome, Sprengel deformity of scapula, crossed fused ectopia (kidneys), Arnold Chiari malformation I | No | Severe | Severe | Deceased | Yes | Yes | No | Pyloric stenosis |
| c.1629delT | p.E543del | 04-0077 | Female | EUR | maternal | Unknown | Unknown | Severe | - | Deceased | Yes | Yes |  | CHD |
| c.1709C>T | p.P570L | 04-0031 | Female | EUR | unknown (singleton) | Father with residual post axial polydactyly | No | Severe | - | Deceased | Yes | No |  | No |
| c.1773G>C | p.E591D | 1511 | Male | EUR | *de novo* | No | No | Mild | None | Alive | No | No | No | No |
| c.1789C>T | p.R597* | 01-0582 | Male | AMR | unknown (singleton) | No | No | None | None | Deceased | No | Yes |  | CHD |
| c.1895-1G>T | p.X632_splice | 1449 | Female | EUR | maternal | Unknown | Unknown | - | - | Deceased | No | No |  |  |
| c.1913C>T | p.T638M | 01-0057 | Female | AMR | *de novo* | Unknown | Unknown | Severe | Moderate | Alive | No | Yes | No | CHD, PH |
| c.1913C>T | p.T638M | 01-0513 | Female | EUR | paternal | No | Affected sibling (N/T), Father with R eventration (+), paternal grandfather with R eventration (+) | - | - | Alive | No | No | No | No |
| c.2122G>A | p.G708S | 04-0025 | Male | EUR | paternal | Father with cleft palate | No | Severe | - | Deceased | Yes | No | Seizures | No |
| c.2122G>A | p.G708S | m1021LEMa | Female | EUR | maternal | No | No | - | - | Deceased | Yes | Yes |  | CHD |
| c.2263C>G | p.R755G | 01-1279 | Male | EUR | paternal<sup>a</sup> | No | Affected sibling (-) | - | - | Deceased | No | No |  | No, bilateral CDH |
| c.2263C>G | p.R755G | 09-0028 | Male | EUR | paternal | Maternal great-aunt with CHD | No | Unk | Unk | Alive | No | Yes | No | CHD |
| c.2461G>C | p.A821P | 03-0008 | Male | EUR | maternal | >3rd degree maternal history: unilateral arm agenesis | No | - | - | Deceased | Yes | No |  | No |
| c.2719dup | p.V907Gfs*73 | 01-0732 | Female | EUR | paternal | No | Paternal aunt with possible CDH (N/T) | Unk | Unk | Alive | Yes | No | No | No |

ECMO = extracorporeal membrane oxygenation, PH = pulmonary hypertension, CHD = congenital heart disease, GU = genitourinary, GI = gastrointestinal
1M PH = pulmonary hypertension status at 1 month, 3M PH = pulmonary hypertension status at third month, – in 1PH and 3PH = deceased before 1 or 3 months
+ positive for familial *LONPI* variant
- negative for familial *LONPI* variant
N/T = not tested for familial *LONPI* variant
<sup>a</sup>cases carried biallelic heterozygous variants

**Figure 4.**
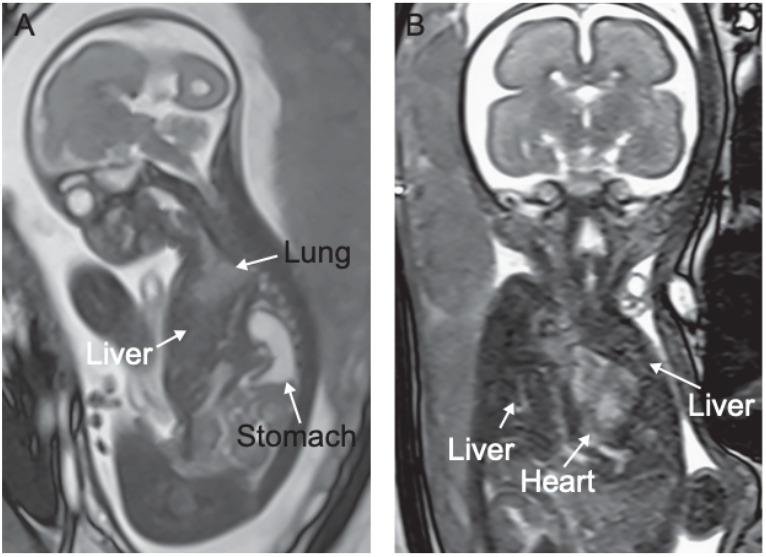
Fetal MRI images of bilateral CDH. (A) Sagittal view shows dorsal herniation of the stomach, ventral herniation of the liver, and anterior displacement of lung remnant. (B) Coronal view shows bilateral herniation of the fetal liver filling both the right and left hemithorax and no lung tissue.

Previous studies reported biallelic variants in *LONP1* in cerebral, ocular, dental, auricular, and skeletal (CODAS) syndrome^53,54^ (MIM: 600373). We compared the locations of the predicted-damaging missense positions in CDH cases and CODAS syndrome cases (Figures 3 and 5). No variants overlap between CDH cases and CODAS syndrome. LONP1 contains three functional domains. CDH damaging variants are concentrated at the core of the domains. Biallelic variants in CODAS syndrome are located on the junction of ATP-binding and proteolytic domains (Figures 3 and 5). The 23 CDH cases with *LONP1* variants didn’t have features of CODAS syndrome.

**Figure 5.**
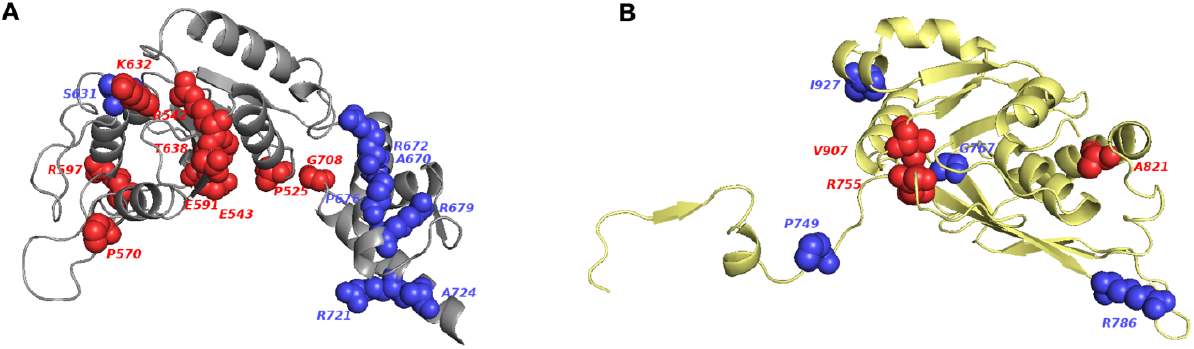
Predicted 3D structure of *LONP1* protein using SWISS-Model. (A) Variants in ATPase domain (gray) of CDH (red) and CODAS (blue). CODAS variants (p.A670-pA724) are clustered at alpha-helix in ATPase domain. (B) Variants in Protease domain (yellow) of CDH (red) and CODAS (blue). CDH variants p.A821, S866 and CODAS variants p.A927 are located at alpha-helix.

### Phenotype of CDH probands with *LONP1* variants

We identified 24 ultra-rare heterozygous variants in 23 sporadic or familial CDH participants (Table 4). The majority (n=17; 73.9%) are of European ancestry and 13 (56.5%) are female (Table 4). Sixteen (70%) were enrolled as neonates. Fourteen of the 23 have a family history of congenital anomalies (Table 4), 6 of whom had a family history of CDH. Nine (39.1%) are complex cases. Six of 9 complex cases have CHD in addition to CDH. We compared the clinical outcomes or phenotypes in CDH cases with *LONP1* damaging variants and other CDH cases (Table 5). Compared to CDH cases without *LONP1* ultra-rare damaging variants, *LONP1* damaging variant carriers are associated with higher neonatal mortality rate prior to initial hospital discharge (69% vs 16%, P=6.4×10^−6^) and greater need for ECMO (56% vs 28%, P=2.3×10^−2^). Compared to CDH cases with other likely damaging variants defined in our previous study^3^, *LONP1* damaging variant carriers had higher neonatal mortality rate prior to discharge (69% vs 24%, P=1.8×10^−3^) and trended towards greater need for ECMO (56% vs 30%, P=0.077).

**Table 5.** *LONP1* deleterious rare variants carriers are associated with higher mortality and need for ECMO.

|  | CDH w/ *LONPI* deleterious variants (n=23) |  |  | CDH w/o *LONPI* deleterious variants (n=806) |  |  | w/ *LONPI* vs. w/o *LONPI* deleterious variants | CDH w/ likely damaging variants (n=98) |  |  | w/ *LONPI* deleterious variants vs. w/ likely damaging variants |
| --- | --- | --- | --- | --- | --- | --- | --- | --- | --- | --- | --- |
|  | Case N | n | % | Control N | n | % | P value | Control N | n | % | P value |
| **Male** | 23 | 10 | 43% | 806 | 477 | 59% | 0.14 | 98 | 47 | 48% | 0.82 |
| **Complex** | 23 | 9 | 39% | 789 | 269 | 34% | 0.66 | 96 | 50 | 52% | 0.35 |
| **Familial CDH** | 19 | 6 | 32% | 806 | 61 | 8% | **2.7E-02** | 98 | 4 | 4% | **1.2E-03** |
| **Neonatal death prior to discharge** | 16 | 11 | 69% | 450 | 72 | 16% | **6.4E-06** | 55 | 13 | 24% | **1.8E-03** |
| **ECMO** | 16 | 9 | 56% | 442 | 124 | 28% | **2.3E-02** | 53 | 16 | 30% | 0.077 |
| **PH at 1m** | 11 | 7 | 64% | 340 | 188 | 55% | 0.76 | 41 | 29 | 71% | 0.72 |
| **PH at 3m** | 6 | 2 | 33% | 260 | 100 | 39% | 1 | 29 | 16 | 55% | 0.4 |

The bold p-values highlight significance. ECMO: extracorporeal membrane oxygenation; PH: pulmonary hypertension

### Inactivation of *Lonp1* in mouse embryonic lung epithelium leads to disrupted lung development and full lethality at birth

The high rate of mortality and need for ECMO in cases with CDH is predominantly due to abnormal lung and pulmonary vascular development causing lung hypoplasia and pulmonary hypertension. Our hypothesis was that impaired or partial loss of *LONP1* function in cases with CDH might contribute directly to abnormal lung development, independent of its role in diaphragm formation. To test this hypothesis, we inactivated *Lonp1* in the embryonic lung epithelium in mice. This was achieved by generating *Shh*^*cre/+*^*;Lonp1*^*fl/fl*^ (hereafter *Lonp1* cKO for conditional knockout) embryos using existing alleles^47^ (International Mouse Strain Resource J:204812). In the mutant the cre recombinase expressed specifically in the epithelium drove *Lonp1* inactivation at the onset of lung initiation (Figure 6A). This led to 100% lethality of the mutants at birth with normal body size (Figures 6B and C). Upon dissection, the mutant lung was composed of large fluid-filled sacs, unlike the controls with normal airways and alveoli (Figure 6D). The lung defect likely contributed to embryonic lethality at birth in these mutant mice.

**Figure 6.**
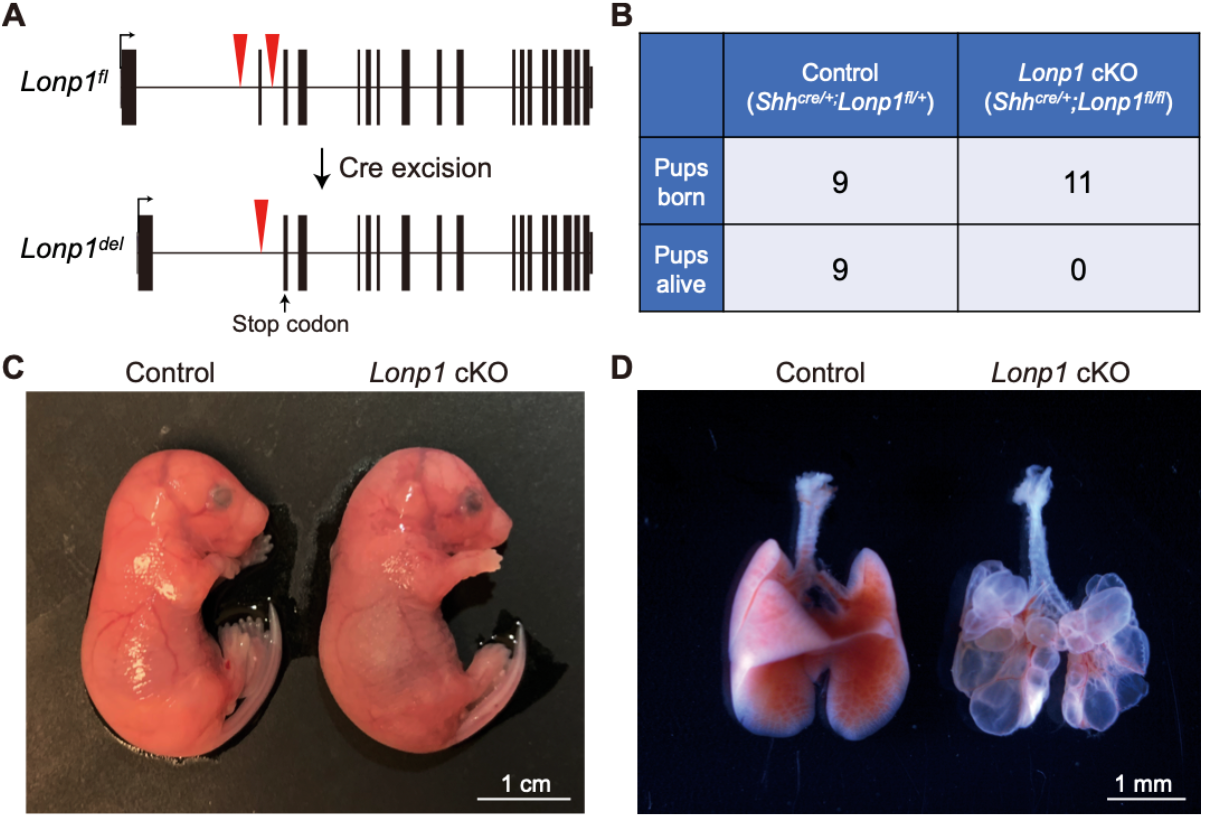
Inactivation of *Lonp1* in mice led to disrupted lung development and lethality at birth. A. Gene structure of mouse *Lonp1*^*fl*^ conditional allele before and after cre-mediated recombination of the *loxP* sites (red triangles). Recombination led to a premature stop codon (arrow) in the second exon. B. Number of embryos genotyped at perinatal stage, showing 100% lethality of the mutant embryos. C. Representative mutant and control embryos at embryonic day (E)18.5, the day of birth. D. Representative mutant and control lungs at E18.5. Scale bars as indicated.

## Discussion

In the current study of 827 CDH trios, we confirmed there is an overall enrichment of damaging *de novo* variants, particularly in constrained genes. We identified *LONP1* and *ALYREF* as novel candidate genes based on enrichment of *de novo* variants. By case-control association, we also confirmed *LONP1* as a genome-wide significant candidate gene contributing to CDH risk through both *de novo* and inherited damaging variants. We demonstrated segregation of the *LONP1* variant with diaphragm defect in five families. We found that CDH individuals with heterozygous ultra-rare damaging variants in *LONP1* have clinical phenotypes frequently including CHD or skeletal anomalies, frequently requiring ECMO, and having a higher mortality than the rest of our CDH cohort. In addition, we confirmed *MYRF* and *ZFPM2* as genes previously associated with CDH^9,14,55,56^. In a mouse model with knock out of *Lonp1* only in the embryonic lung epithelium with an intact diaphragm, we demonstrated reduced pulmonary growth and branching, resulting in perinatal lethality that suggests that the higher mortality rate and need for ECMO in human is due to a primary effect of *LONP1* on pulmonary development in addition to diaphragm development.

The burden of damaging *de novo* variants in CDH is consistent with previous studies^9,14,15^, and damaging *de novo* variants are more frequent in complex CDH compared to isolated CDH cases. Similar patterns have been observed in complex congenital heart disease with other congenital anomalies or neurodevelopmental disorders compared with isolated congenital heart disease^50^ and autism with/without intellectual disability^57^. Deleterious *de novo* variants are more frequent in many severe early-onset diseases with reduced reproductive fitness compared to the general population^58^. The higher frequency of *de novo* LGD variants in female relative to male CDH cases supports the “female protective model” similar to autism^52,59,60^, which means that risk variants have larger effects in males than in females so that females require a higher burden to reach the same diagnostic threshold as males.

Both *de novo* and rare inherited variant analyses highlight *LONP1* as a novel CDH candidate gene. Approximately 3% of individuals in our CDH cohort are heterozygous for *LONP1* rare variants. Three variants (p. T638M, p.G708S and p.R755G) are recurrently and independently found in unrelated families. CDH cases with *LONP1* variants had higher mortality in the neonatal period compared with other CDH cases. Biallelic variants in *LONP1* have been reported in CODAS, a multi-system developmental disorder characterized by cerebral, ocular, dental, auricular, and skeletal anomalies^61^. The Lonp1 holoenzyme is a homohexamer with six identical subunits. Each subunit consists of a mitochondrial-targeting sequence (MTS), a substrate recognition and binding (N) domain, an ATPase (AAA+) domain, and a proteolytic (P) domain. Biallelic missense variants reported in CODAS individuals are mostly located in the junction of ATP-binding and proteolytic domains of LONP1 while the heterozygous variants identified in CDH individuals are located in the main domains of LONP1. Notably, there are no overlapping variants between CDH and CODAS individuals. Most of the variants in CODAS are located in the alpha-helix and may affect the interactions of subunits^61^. Variants in CDH may interrupt the proteolytic and ATP binding domains, resulting in the dysfunction of LONP1. Homozygous deletion of LONP1 in mice is embryonic lethal, due to progressive loss of mtDNA with subsequent failure to meet energy requirements for embryonic development^62^. Heterozygous *Lonp1*^*+/-*^ mice develop normally without obvious abnormalities, but lonp1 expression decreased in both RNA and protein levels^62^. Analysis of Lonp1 expression in heterozygous mice indicated a 50% reduction at both RNA and protein levels in these animals. These data suggest different mechanisms of *LONP1* in diseases with biallelic and monoallelic variants. Of note, one CDH individual carried biallelic variants (p.P525L and p.R755G). No additional phenotypes were noted, perhaps because the baby died at 8-9 hours after birth with severe bilateral CDH (Figure 4).

Lonp1 is a nuclear-encoded mitochondrial protease. Besides binding of mtDNA^63^, Lonp1 was discovered as an ATP-dependent protease involved in the degradation of misfolded or damaged proteins^64-66^. Accumulation of misfolded proteins has been observed in the impaired lungs of developing mice with deletion of other ATP-dependent proteins^67^. The immature lung development and neonatal respiratory failure of our *Lonp1* cKO mice could be due to the inactivation of Lon protease, which results in the accumulation of misfolded proteins and activation of the unfolded protein response (UPR) pathway^68^. UPR activation during development could lead to reduced cell proliferation and cause other congenital anomalies including congenital heart disease^69^.

Lonp1 also acts as a chaperone that interacts with other mitochondrial proteins to regulate several cellular processes^70^. Lon expression may stimulate cell proliferation^71^ and Lon downregulation may impair mitochondrial structure and function and cause apoptosis^72,73^. Alterations in cell proliferation, differentiation and migration can all lead to CDH. Myogenic cell differentiation and migration are essential during formation of the diaphragm^74^. Myogenic differentiation requires increased expression of mitochondrial biogenesis-related genes including Lon^75^. The variants could cause an increased probability of failure of myogenesis during embryonic development, consequently resulting in the hernia.

The neonatal mortality of probands with *LONP1* deleterious variants is much higher than CDH neonates without *LONP1* deleterious variants or CDH neonates with likely damaging variants in genes other than *LONP1*. CDH neonates with *LONP1* deleterious variants frequently required ECMO. In mice with *Lonp1* knock out at the onset of lung development, 100% newborn pups died shortly after birth, with severe pulmonary defects. Thus, *LONP1* could represent a class of CDH genes with high mortality due to primary developmental effects on the lung, resulting in more severe pulmonary defects than would occur secondary to lung compression by herniated abdominal viscera alone. This suggests that we should try to differentiate primary from secondary developmental effects on the lung as we phenotype newborns with CDH and as we investigate the mechanisms action of CDH candidate genes.

The RNA-binding protein ALYREF plays a key role in nuclear export through binding to the 5’ and the 3’ regions of mRNA^76,77^. It acts as an RNA 5-methylcytosine (m^5^C) adaptor to regulate the m^5^C modification^78,79^. Disruption of ALYREF could affect the m^5^C modification, resulting in abnormal cell proliferation and migration^79^. Previous studies^50^ identified several RNA binding proteins (RBPs) playing essential roles in autism and congenital birth defects including CHD. RBFOX2, an RBP that regulates alternative splicing, is critical for zebrafish heart development^80^ and *de novo* variants in *RBFOX2* are associated with congenital heart defects^50^. Dozens of RBPs have established roles in autism spectrum disorder. RBFOX1^81,82^, an RNA splicing factor, regulates expression of large genetic networks during early neuronal development including autism. The other RBPs such as FMRP^83^, CELF4, CELF6^84^, have also been implicated in autism. As an RBP, ALYREF may play a similar role in congenital anomalies and neurodevelopmental disorders. Two *de novo* LGDs in *ALYREF* were identified in our CDH cohort. One had an isolated CDH and the other had CDH and a ventricular septal defect. Similarly, two CDH cases carried *de novo* variants in *SYMPK*, another RBP identified with FDR<0.1 in extTADA. One had a *de novo* predicted deleterious missense variant and isolated CDH and the other had a *de novo* LGD with complex CDH with congenital heart disease, central nervous system anomaly, and genitourinary anomaly.

We found further support for the previously reported CDH genes *ZFPM2* and *MYRF*. We have identified six ultra-rare LGD variants in *ZFPM2* in our CDH cohort, accounting for 0.7% of our cases (Figure S6). Three were complex cases, all with minor cardiac malformations. Specifically, two females had atrial septal defects and 1 male had an enlarged aortic root. The other three heterozygotes had isolated CDH. *ZFPM2* is expressed in the septum transversum of the diaphragm during early development, and Fog2^-/-^ mice generated through chemical mutagenesis have been shown to have diaphragmatic eventration and pulmonary hypoplasia^55^. *ZFPM2* physically interacts with NR2F2^85^ and GATA4^86^, two other components of the retinoid signaling pathway implicated in diaphragm and lung development^87^. Our results further support the pleiotropic role of *ZFPM2* in the development of CDH.

*MYRF* was implicated in our previous *de novo* variant report^9^ as a gene for cardiac-urogenital syndrome (MIM: 618280), and we identified one more additional *de novo* variant in this cohort (Figure S7). There are now more than 10 variants implicated in CDH with additional anomalies (HGMD® professional 2021.1). *MYRF* is highly expressed in epithelial cells. Diaphragm is composed of epithelial-like mesothelial cells derived from the mesoderm of the pleuroperitoneal folds (PPFs) through cell proliferation, migration, and epithelial-to-mesenchymal transition^88^. Single cell analysis^89^ in fetal gonads suggests the cells that highly express *MYRF* also express *WT1* and *NR2F2*, two genes associated with diaphragmatic hernia. Previously, we also demonstrated^9^ that individuals with pathogenic variants in *MYRF* have decreased expression of *GATA4. WT1, NR2F2* and *GATA4* are all important in RA signaling in the developing diaphragm^1^. Therefore, the damaging variants in *MYRF* may affect the RA signaling pathway, leading to diaphragmatic hernia and other anomalies.

Among the 734 CDH trios with WGS data, we identified a total of 87 *de novo* CNVs and 4 of them are recurrent genes or CNVs. Given the rarity of *de novo* CNVs and small sample size, there were limited data to analyze the differential burden between cases and controls in this study. Future studies with larger sample sizes will improve the power to analyze CNVs and structural variants in CDH.

In summary, our analysis of *de novo* and ultra-rare inherited variants identified two new CDH candidate genes *LONP1* and *ALYREF* and confirmed previous associations of *MYRF* and *ZFPM2* with CDH. The identification of specific highly risk genes would enhance prenatal or early postnatal counseling and decision making, especially with rapid turnaround of WGS or exome sequencing results. It is likely that transmitted rare variants also contribute to other cases in our cohort, but we require a larger sample size to identify these genes confidently. Future studies will also leverage data from other developmental disorders and integrating genomic data during development.

## Data Availability

All public data from databases and studies were described in the manuscript. The genetic data is accessible through the Diaphragmatic Hernia Research & Exploration; Advancing Molecular Science study (DHREAMS, http://www.cdhgenetics.com/) after necessary clearances; please contact author Wendy K. Chung for more details. The SPARK dataset used in this analysis is preparing to publish and has been given to the Simons Foundation Autism Research Initiative (SRARI) for public distribution. Scientists wishing to access the data set can do so through application to SFARI. The WHICAP dataset analyzed during the current study are available via contact with Badri N. Vardarajan.

## Supplemental Data

Supplemental Data include notes, 7 figures and 7 tables.

## Acknowledgements

We would like to thank the patients and their families for their generous contribution. We are grateful for the technical assistance provided by Na Zhu, Patricia Lanzano, Jiangyuan Hu, Jiancheng Guo, Suying Bao, Charles LeDuc, Liyong Deng, Donna Garey, and Anketil Abreu from Columbia University, Jennifer Lyu at Boston Children’ s Hospital, and Caroline Coletti at Massachusetts General Hospital. We thank our clinical coordinators across the DHREAMS centers: Jessica Conway at Washington University School of Medicine, Melissa Reed, Elizabeth Erickson, and Madeline Peters at Cincinnati Children’s Hospital, Sheila Horak and Evan Roberts at Children’s Hospital & Medical Center of Omaha, Jeannie Kreutzman and Irene St. Charles at CS Mott Children’s Hospital, Tracy Perry at Monroe Carell Jr. Children’s Hospital, Dr. Michelle Kallis at Northwell Health, Andrew Mason and Alicia McIntire at Oregon Health and Science University, Gentry Wools and Lorrie Burkhalter at Children’s Medical Center Dallas, Elizabeth Jehle at Hassenfeld Children’s Hospital, Michelle Knezevich and Cheryl Kornberg at Medical College of Wisconsin, Min Shi at Children’s Hospital of Pittsburgh. We would also like to acknowledge Terry Buchmiller at Boston Children’s Hospital, and the other pediatric surgeons and clinicians who referred patients to our studies.

The whole genome sequencing data were generated through NIH Gabriella Miller Kids First Pediatric Research Program (X01HL132366, X01HL136998, X01HL155060). This work was supported by NIH grants R01HD057036 (L.Y., J.W., W.K.C.), R03HL138352 (A.K., W.K.C., Y.S.), R01GM120609 (H.Q., Y.S.), UL1 RR024156 (W.K.C.) 1P01HD068250 (P.K.D, F.A.H., J.M.W., W.K.C., Y.S., J.M.Z, D.J.M, X.S.) and NSFC81501295 (L.Y.). Additional funding support was provided by grants from CHERUBS, CDHUK, and the National Greek Orthodox Ladies Philoptochos Society, Inc. and generous donations from the Williams Family, Wheeler Foundation, Vanech Family Foundation, Larsen Family, Wilke Family and many other families. Whole genome sequencing data can be obtained from dbGAP through accession phs001110. WHICAP study is supported by funding from NIA RF1AG054023 (B.N.V.). Biogen Inc provided support for whole-exome sequencing for the WHICAP cohort.

## Web Resources

DHREAMS study, http://www.cdhgenetics.com/

Integrative Genome Viewer (IGV), http://software.broadinstitute.org/software/igv

ClinGen genome dosage map, https://dosage.clinicalgenome.org

DECIPHER, https://www.deciphergenomics.org

Combined Annotation Dependent Depletion (CADD), https://cadd.gs.washington.edu/

GenBank, https://www.ncbi.nlm.nih.gov/genbank/

Genome Aggregation Database (gnomAD), https://gnomad.broadinstitute.org/

Online Mendelian Inheritance in Man (OMIM), https://www.omim.org/

PyMOL molecular viewer, https://pymol.org/2/

Mouse Genome Informatics (MGI), http://www.informatics.jax.org

The Human Protein Atlas, https://www.proteinatlas.org/

