## Supplemental Data for "Rare and *de novo* variants in 827 congenital diaphragmatic hernia probands implicate *LONP1* and *ALYREF* as new candidate risk genes"

**Supplemental Materials and Methods**

*extTADA analysis.* To estimate yield insight into the genetics of CDH, we performed an empirical Bayesian model of rare-variant genetic architecture, extTADA[1] (Extended Transmission and *de novo* Association) based on burden of *de novo* variants. The extTADA model is developed based on a previous integrated empirical Bayesian model TADA[2] and estimate mean effect sizes and risk-gene proportions from the genetic data by estimating parameters using MCMC (Markov Chain Monte Carlo) process.

Two major parameters, relative risk of a gene causing a disease $\gamma$ and the proportion of disease risk genes across the local gene groups $\pi$, were estimated by the connection to variant fold enrichment (FE), which was calculated as the number of observed variants divided by the number expected. The expectation was from the baseline mutation rate of each gene and was the same as used in burden analysis. Assuming the background mutation rate for each gene is $\mu$, total number of genes in the gene set is *m* and total number of sequenced samples is *N*, then the observed variants were: $X = \pi m \times 2\gamma\mu N + (1-\pi)m \times 2\mu N$. The expected variants were: $X_e = \pi m \times 2\mu N + (1-\pi)m \times 2\mu N$.

FE can be calculated from the data, parameters $\gamma$, $\pi$ and $\beta$ are estimated accordingly using a Hamiltonian Monte Carlo (HMC) MCMC method implemented in the "rstan" package (Carpenter et al., 2017): $FE = \frac{X}{X_e} = \pi(\gamma - 1) + 1$, where $\gamma \sim Gamma\ (\bar{\gamma}\beta, \beta)$.

Bayes factors can be estimated as following: $B = \frac{P(X|H1)}{P(X|H0)} \sim \frac{Pois(2\gamma\mu N)}{Pois(2\mu N)}$, where H0 is null hypothesis and H1 is alternative hypothesis, $\gamma$=1.

From Bayes' theorem, the posterior odds are equal to the Bayes factor times the prior odds: $\frac{P(H1|X)}{P(H0|X)} = \frac{P(X|H1)}{P(X|H0)} \times \frac{P(H1)}{P(H0)}$, where $P(H1)$ is estimated $\pi$ and $P(H0)$ is (1-$\pi$).

Assuming the posterior probability of association (PPA), $P(H1|X)$is $q$, so the posterior probability of the null model $P(H0|X)$is $q_0 = (1-q)$, where $q = \frac{B\pi}{1-\pi+B\pi}$. Per-gene based FDR is calculated from $q_0$ as the following: $q_0$ is ranked in an increasing order for all the genes, then FDR is the sum of total $q_0$ smaller than the current rank *k* divided by the total number of genes *k* with smaller $q_0$.

To inform the parameter estimation with prior knowledge, we stratify the whole genome genes into constrained genes (ExAC pLI score ≥0.5) and non-constrained genes (other genes). The extTADA model was applied to each group of genes to estimate the local parameters and calculate PPAs. Then we combined the PPAs of 2 groups together to calculate a final genome-wide FDR (false discovery rate).

### Supplemental Figures

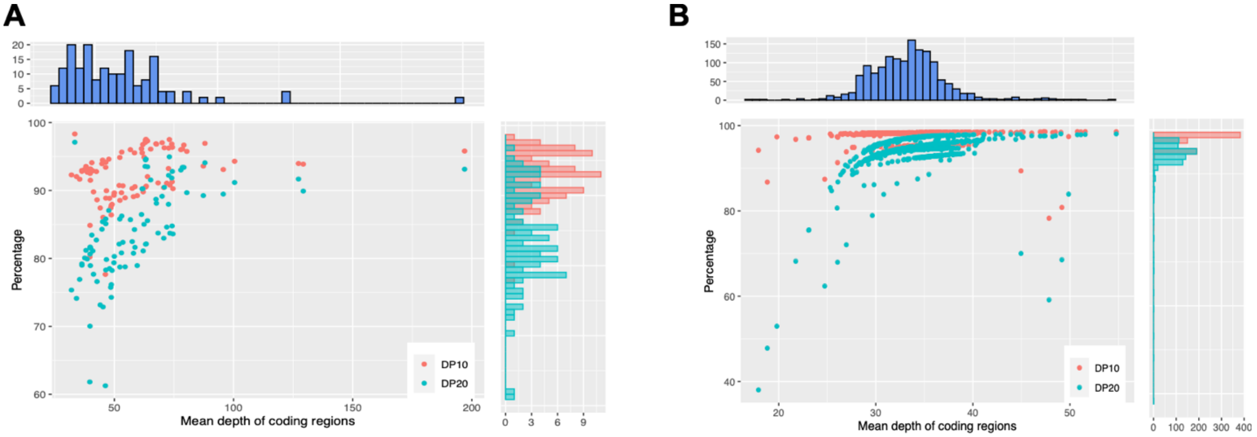


**Figure S1. Depth of coverage for all cases across all targeted regions.** DP10/DP20 is the percentage of targeted regions that are covered by at least 10/20 unique reads. Scatter plot and marginal histograms for mean depth and DP10/DP20 are shown for (A) 96 exome sequencing cases and (B) 735 whole-genome sequencing cases. Coverage statistics were calculated on the coding regions (including canonical splice sites) of GENCODE genes (v19).

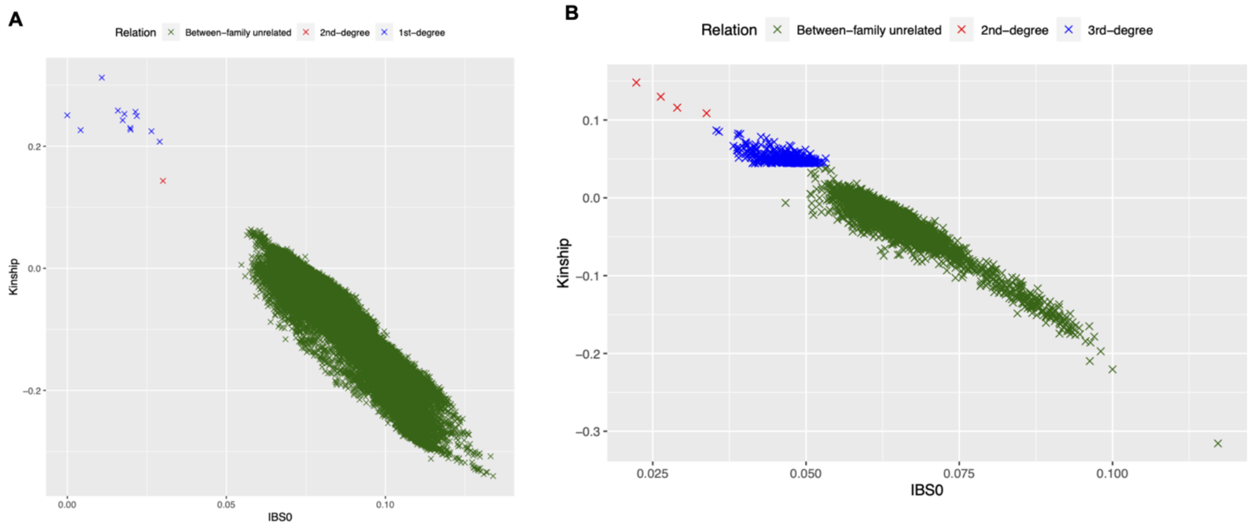


**Figure S2. Relatedness predicted by KING.** (A) CDH cases; (B) controls

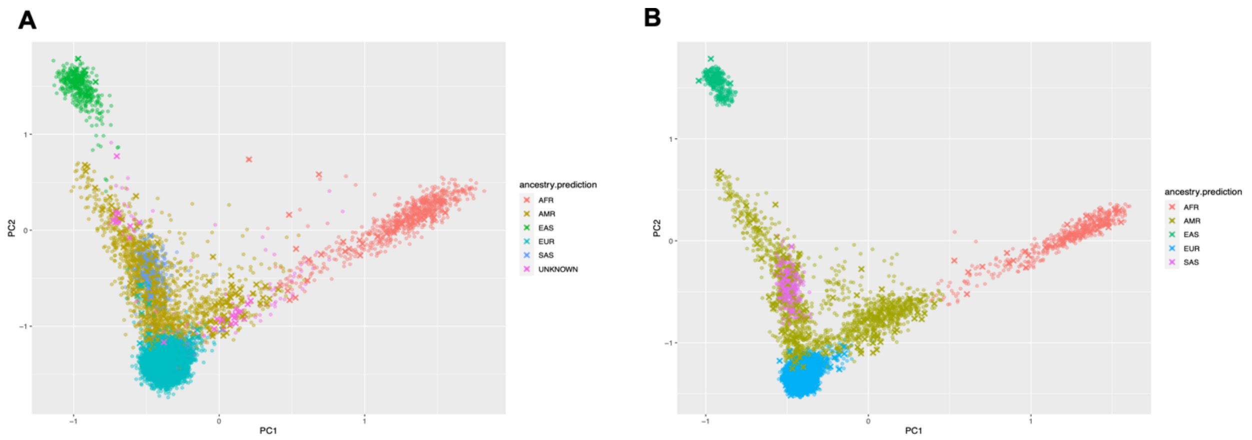


**Figure S3. Principle components analysis (PCA) of ethnicity using peddy.** Cross signals are cases and dots are controls. (A) all samples, (B) samples after matching-ancestry for case-control analysis. AFR: African; AMR: Admixed American/Latinx; EAS: East Asian; EUR: European; SAS: South Asian

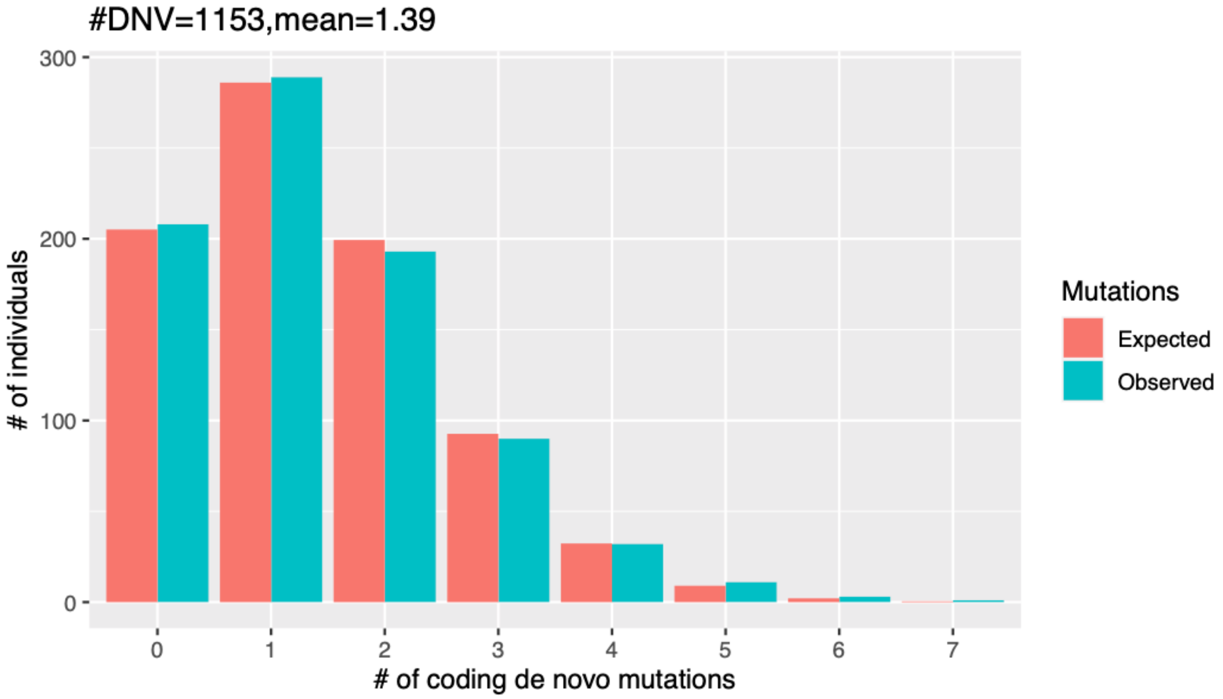


**Figure S4. Distribution of de novo coding variants per proband.** The observed number of variants (green) were compared with the expectation (red) assuming a Poisson distribution with mean equals to the average number of variants per proband.

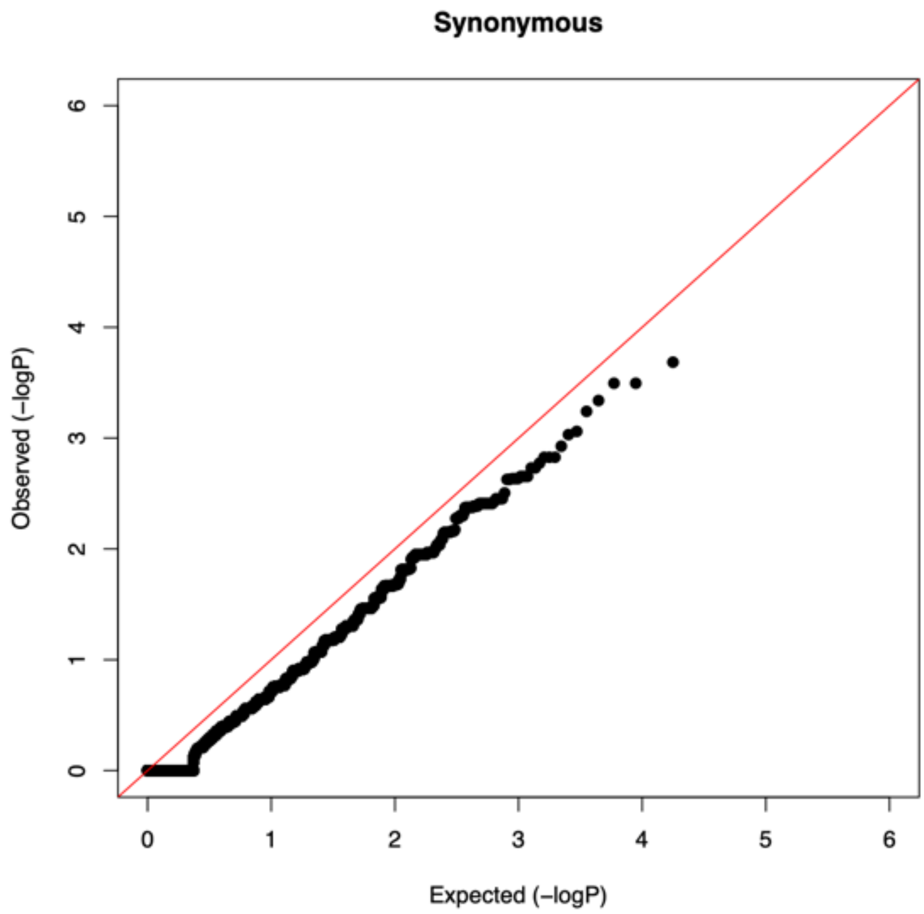


**Figure S5. Gene-level burden test of ultra-rare synonymous.**

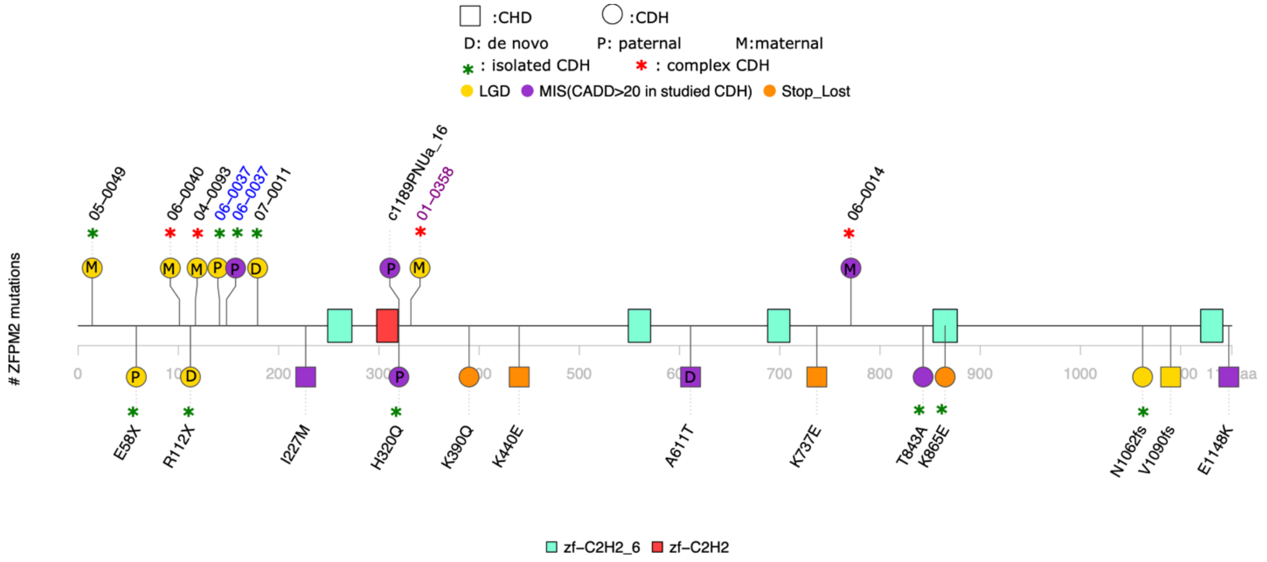


**Figure S6. Mutation locations in *ZFPM2* of CDH and published CDH/CHD cases.** There are two main domains in ZFPM2, zf-C2H2_6 domain and zf-C2H2 domain. Positions indicated at upper structure are mutations carriers' IDs in CDH, the colored sample IDs are familiar CDH cases and one color is in one family. Deleterious heterozygous variant such as LGD and missense with CADD $\geq$20 and minor allele frequency (MAF) <1e-5 alleles in CDH were presented. Deleterious missense is presented in purple, LGD in yellow, stop loss in orange. Inheritance pattern were labelled in circles of mutations. Complex or isolated CDH are presented for each CDH case. Positions at lower structure are mutations in published CDH/CHD samples[3-6].

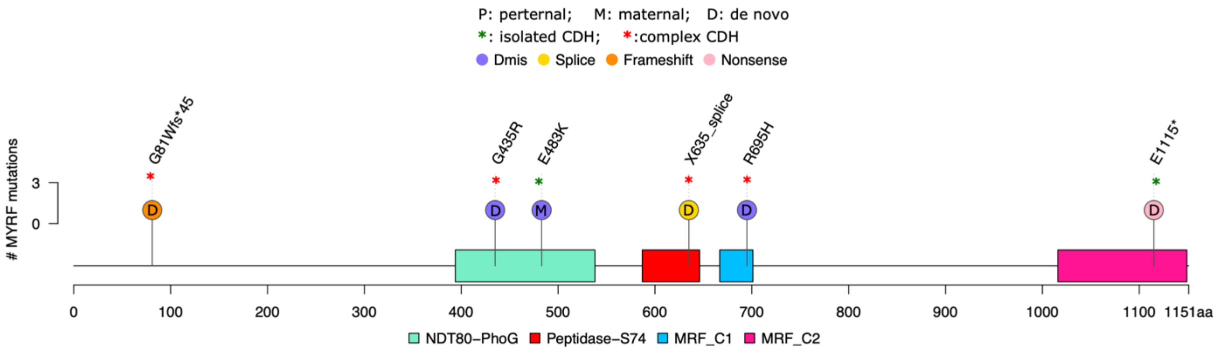


**Figure S7. Mutation locations in *MYRF* of CDH.** There are four main domains in MYRF, NDT80/PhoG like DNA-binding family (NDT80_PhoG), chaperone of endosialidase (Peptidase_S74), myelin gene regulatory factor-C-terminal domain 1 (MRF_C1) and myelin gene regulatory factor-C-terminal domain 2 (MRF_C2). Deleterious heterozygous variant such as LGD and missense with CADD ≥27 and minor allele frequency (MAF) <1e-5 alleles in CDH were presented. Deleterious missense is presented in purple, splice in yellow, frameshift in orange and nonsense in pink. Inheritance pattern were labelled in circles of mutations. Complex or isolated CDH are presented for each CDH case.

### Supplemental Tables

#### Table S1a. Sequencing batches of 827 studied CDH

<table>
<tr><th>Data Type</th><th>Seq Center</th><th>Protocol</th><th>#Studied</th><th>#Published</th></tr>
<tr><td rowspan="4">Exome</td><td>CUGC/NYGC</td><td>SureSelect V2/V4</td><td>27</td><td>17</td></tr>
<tr><td>University of Washington</td><td>SeqCap EZ V2</td><td>21</td><td>20</td></tr>
<tr><td>CloudHealth Genomics, China</td><td>SeqCap EZ V3</td><td>11</td><td>2</td></tr>
<tr><td>University of Washington</td><td>VCRome V2</td><td>34</td><td>34</td></tr>
<tr><td rowspan="4">Genome</td><td>Baylor College of Medicine</td><td>PCR-free lib prep</td><td>191</td><td>190</td></tr>
<tr><td>Broad Institute</td><td>PCR-free lib prep</td><td>310</td><td>255</td></tr>
<tr><td rowspan="2">Broad Institute</td><td>PCR-free lib prep</td><td>217</td><td>54</td></tr>
<tr><td>PCR-plus lib prep</td><td>16</td><td>2</td></tr>
<tr><td>Total</td><td></td><td></td><td>827</td><td>574</td></tr>
</table>

CUGC: Columbia University Genome Center; NYGC: New York Genome Center

#### Table S1b. Enrollment centers of 827 studied CDH

| Site_No in REDCap | Site | #Studied |
|---|---|---|
| 1 | Columbia | 360 |
| 2 | Washington University | 44 |
| 3 | University of Pittsburgh | 12 |
| 4 | University of Cincinnati | 74 |
| 5 | Omaha Children's Hospital / University of Nebraska | 61 |
| 6 | University of Michigan | 45 |
| 7 | Vanderbilt University | 29 |
| 9 | Wisconsin Children's Hospital | 29 |
| 10 | Cairo University | 24 |
| 11 | North Shore LIJ | 5 |
| 12 | Oregon | 12 |
| 14 | Dallas | 11 |
| 15 | Poland | 17 |
| 16 | U of Wisconsin, Madison | 4 |
| 19 | MGH Boston | 100 |
| Total | | 827 |

### Table S3. Burden of de novo coding variants.

| | All | | | | Female | | | | Male | | | | Complex | | | | Isolated | | | |
|---|---|---|---|---|---|---|---|---|---|---|---|---|---|---|---|---|---|---|---|---|
| | **#Obs** | **#Exp** | **RR** | **P** | **#Obs** | **#Exp** | **RR** | **P** | **#Obs** | **#Exp** | **RR** | **P** | **#Obs** | **#Exp** | **RR** | **P** | **#Obs** | **#Exp** | **RR** | **P** |
| **All genes** | | | | | | | | | | | | | | | | | | | | |
| Syn | 247 | 273 | 0.9 | 0.12 | 105 | 113 | 0.9 | 0.48 | 142 | 160 | 0.89 | 0.17 | 78 | 92 | 0.85 | 0.17 | 164 | 174 | 0.9 | 0.45 |
| Mis | 694 | 609 | 1.1 | **0.00071** | 284 | 252 | 1.1 | **0.047** | 410 | 357 | 1.15 | **0.0054** | 241 | 204 | 1.18 | **0.012** | 443 | 389 | 1.1 | **0.0066** |
| D-mis | 292 | 220 | 1.3 | **3.10E-06** | 133 | 91 | 1.5 | **3.20E-05** | 159 | 129 | 1.23 | **0.0093** | 108 | 74 | 1.47 | **0.00018** | 180 | 140 | 1.3 | **0.0011** |
| LGD | 126 | 85 | 1.5 | **3.60E-05** | 58 | 35 | 1.6 | **0.00038** | 68 | 50 | 1.36 | **0.013** | 52 | 29 | 1.82 | **7.20E-05** | 72 | 54 | 1.3 | **0.021** |
| **Constrained genes** | | | | | | | | | | | | | | | | | | | | |
| Syn | 92 | 93 | 0.99 | 0.96 | 41 | 39 | 1.1 | 0.75 | 51 | 54 | 0.94 | 0.73 | 36 | 31 | 1.2 | 0.37 | 56 | 60 | 0.9 | 0.7 |
| Mis | 263 | 208 | 1.26 | **0.00027** | 111 | 87 | 1.3 | **0.014** | 152 | 121 | 1.25 | **0.0064** | 89 | 70 | 1.3 | **0.027** | 167 | 133 | 1.3 | **0.0042** |
| D-mis | 139 | 88 | 1.59 | **3.40E-07** | 59 | 36 | 1.6 | **0.00063** | 80 | 51 | 1.57 | **0.00015** | 52 | 29 | 1.8 | **0.00014** | 85 | 56 | 1.5 | **0.00028** |
| LGD | 65 | 30 | 2.2 | **1.80E-08** | 37 | 12 | 3 | **1.20E-08** | 28 | 17 | 1.63 | **0.015** | 31 | 10 | 3.1 | **6.70E-08** | 33 | 19 | 1.8 | **0.0026** |
| **Non-constrained genes** | | | | | | | | | | | | | | | | | | | | |
| Syn | 155 | 180 | 0.86 | 0.062 | 64 | 74 | 0.9 | 0.27 | 91 | 106 | 0.86 | 0.16 | 42 | 60 | 0.7 | 0.017 | 108 | 115 | 0.9 | 0.58 |
| Mis | 431 | 401 | 1.08 | 0.13 | 173 | 165 | 1.1 | 0.53 | 258 | 235 | 1.1 | 0.14 | 152 | 134 | 1.13 | 0.13 | 276 | 256 | 1.1 | 0.2 |
| D-mis | 153 | 132 | 1.16 | 0.074 | 74 | 54 | 1.4 | **0.0098** | 79 | 78 | 1.02 | 0.86 | 56 | 44 | 1.26 | 0.083 | 95 | 84 | 1.1 | 0.25 |
| LGD | 61 | 56 | 1.1 | 0.46 | 21 | 23 | 0.9 | 0.83 | 40 | 33 | 1.22 | 0.19 | 21 | 19 | 1.13 | 0.56 | 39 | 35 | 1.1 | 0.56 |

#Obs: number of observed variants; #Exp: number of expected variants; RR: relative risk; Constrained genes: genes with ExAC pLI>0.5

**Table S4. The proportion of risk genes and mean relative risk for CDH cases using extTADA.**

| | Proportion of risk genes | Relative risk (D-mis) | Relative risk (LGD) |
|---|---|---|---|
| All genes | 0.007 | 11.35 | 23.97 |
| Constrained genes | 0.037 | 10.01 | 18.30 |
| Non-constrained genes | 0.006 | 3.81 | 5.24 |

**Table S6. Sample size for Population-based case-control analysis. EUR: European; AMR: Admixed American/Latinx; AFR: African; EAS: East Asian; SAS: South Asian**

| | CDH Case (n=748) | | SPARK control(n=13,369) | | WHICAP control | | controls (case*15=11,220) | |
|---|---|---|---|---|---|---|---|---|
| | Sample size | Ethnic proportion | Sample size | Ethnic proportion | | #Control/#Case | Sample size | Ethnic proportion |
| EUR | 576 | 77.01% | 10,513 | 79.64% | | 18.2 | 8640 | 77.01% |
| AMR | 110 | 14.71% | 1594 | 11.36% | 908 | 22.7 | 1650 | 14.71% |
| AFR | 31 | 4.14% | 646 | 4.61% | | 20.8 | 465 | 4.14% |
| EAS | 15 | 2.01% | 368 | 2.62% | | 24.5 | 225 | 2.01% |
| SAS | 16 | 2.14% | 248 | 1.77% | | 15.5 | 240 | 2.14% |

### Table S7. Similar frequency of ultra-rare variants among all and each ethnic cases and controls.

| | Variant class | Case | Control | Rate in case | Rate in control | Enrichment | P |
|---|---|---|---|---|---|---|---|
| All (748 cases vs 11,220 controls) | SYN | 13227 | 198396 | 17.68 | 17.68 | 1 | 1 |
| | Inframe | 738 | 11093 | 0.9866 | 0.9887 | 1 | 0.97 |
| | LGD | 3427 | 55530 | 4.582 | 4.949 | 0.93 | 9.70E-06 |
| | MIS | 29408 | 441964 | 39.32 | 39.39 | 1 | 0.75 |
| | SNV | 44387 | 667484 | 59.34 | 59.49 | 1 | 0.61 |
| | Indels | 2612 | 43118 | 3.492 | 3.843 | 0.91 | 1.50E-06 |
| AFR (31 cases vs 465 controls) | SYN | 627 | 9408 | 20.23 | 20.23 | 1 | 1 |
| | Inframe | 42 | 630 | 1.355 | 1.355 | 1 | 1 |
| | LGD | 153 | 2996 | 4.935 | 6.443 | 0.77 | 0.001 |
| | MIS | 1302 | 19199 | 42 | 41.29 | 1 | 0.54 |
| | SNV | 2003 | 29632 | 64.61 | 63.72 | 1 | 0.55 |
| | Indels | 128 | 2769 | 4.129 | 5.955 | 0.69 | 2.30E-05 |
| Hispanic (110 cases vs 1650 controls) | SYN | 2549 | 38213 | 23.2 | 23.2 | 1 | 0.98 |
| | Inframe | 123 | 1842 | 1.12 | 1.12 | 1 | 0.96 |
| | LGD | 572 | 10506 | 5.2 | 6.37 | 0.82 | 1.40E-06 |
| | MIS | 5284 | 84751 | 48 | 51.4 | 0.94 | 1.90E-06 |
| | SNV | 8120 | 128930 | 73.8 | 78.1 | 0.94 | 5.60E-07 |
| | Indels | 438 | 7149 | 3.98 | 4.33 | 0.92 | 0.088 |
| EUR (576 cases vs 8,640 controls) | SYN | 8953 | 134306 | 15.5 | 15.5 | 1 | 1 |
| | Inframe | 499 | 7511 | 0.866 | 0.869 | 1 | 0.96 |
| | LGD | 2476 | 37800 | 4.3 | 4.38 | 0.98 | 0.4 |
| | MIS | 20582 | 305088 | 35.7 | 35.3 | 1 | 0.1 |
| | SNV | 30821 | 458052 | 53.5 | 53 | 1 | 0.12 |
| | Indels | 1832 | 29068 | 3.18 | 3.36 | 0.95 | 0.019 |
| EAS (15 cases vs 225 controls) | SYN | 575 | 8624 | 38.33 | 38.33 | 1 | 1 |
| | Inframe | 32 | 479 | 2.133 | 2.129 | 1 | 1 |
| | LGD | 94 | 1907 | 6.267 | 8.476 | 0.74 | 0.0036 |
| | MIS | 1116 | 16762 | 74.4 | 74.5 | 1 | 0.99 |
| | SNV | 1740 | 26135 | 116 | 116.2 | 1 | 0.97 |
| | Indels | 85 | 1753 | 5.667 | 7.791 | 0.73 | 0.0032 |
| SAS (16 cases vs 240 controls) | SYN | 523 | 7845 | 32.7 | 32.7 | 1 | 1 |
| | Inframe | 42 | 631 | 2.62 | 2.63 | 1 | 1 |
| | LGD | 132 | 2321 | 8.25 | 9.67 | 0.85 | 0.08 |
| | MIS | 1124 | 16164 | 70.2 | 67.3 | 1 | 0.17 |
| | SNV | 1703 | 24735 | 106 | 103 | 1 | 0.2 |
| | Indels | 129 | 2379 | 8.06 | 9.91 | 0.81 | 0.021 |
